# Evaluating and optimizing COVID-19 vaccination policies: a case study of Sweden

**DOI:** 10.1101/2021.04.07.21255026

**Authors:** Henrik Sjödin, Joacim Rocklöv, Tom Britton

## Abstract

We evaluate the efficiency of vaccination scenarios for COVID-19 by analysing a data-driven mathematical model. Healthcare demand and incidence are investigated for different scenarios of transmission and vaccination schemes. Our results suggest that reducing the transmission rate affected by invading virus strains, seasonality and the level of prevention, is most important. Second to this is timely vaccine deliveries and expeditious vaccination management. Postponing vaccination of antibody-positive individuals reduces also the disease burden, and once risk groups have been vaccinated, it is best to continue vaccinating in a descending age order.

## Introduction

The COVID-19 pandemic has resulted in millions of deaths and seriously ill people, affecting societies all over the world. Fortunately, effective vaccines (1,2) have now become available and are being distributed in many countries. At the same time new and more transmissible and virulent strains invade. Perhaps most notable, the new lineage B.1.1.7 originating from the UK, which has become increasingly dominant in Europe and Sweden and estimated to have a 39-130% higher reproduction number (3), and has been suggested to cause more severe illness (4). As a consequence, it is now of paramount importance to roll out the vaccines as quickly and effectively as possible in order to minimize the damage from new epidemic waves caused by the circulation of new virus strains to which current control measures are inadequate. Considering the time required for implementation of vaccination to sufficient coverage of the adult population, the vaccine strategy can make a direct difference for the number of people becoming severely ill or dying in COVID-19 in the coming months.

The focus of the present paper is to describe and evaluate scenarios and vaccination strategies with respect to incidence and health care load from COVID-19 in countries similar to Sweden up to 1 October 2021. The factors of the different scenarios include vaccination-order policies, vaccine-delivery delays, vaccination implementation rate and transmission rate. The latter in turn is affected by the mean transmissibility of invading virus strains, seasonality, and the overall effect of intervention measures.

## Methodology

We develop and use an epidemic SEIR model with 10-year age-strata and structure alike Sweden, which provides estimates of incidence, hospital care occupancy, critical care occupancy, fatalities, and individuals recovered from natural infection or protected by vaccine. A detailed description of the model and parameters is given in the S.M. Here we just give a brief description of the model which is an extension of a previously established model (5). The model uses the age structure of the Swedish population in 2019, and contact rates between 10-year age-groups also stemming from Sweden (6). The model parameters for health-care demand are derived from Swedish data (7, 8, see S.M.), while some transmission parameters are based on international data (9). The model is run from December 1, 2020 until October 1, 2021. The initial conditions (on December 1) reflect data from Sweden (7, 8, see S.M.). The transmission rate follows the estimated daily *R*_*t*_ from the Public Health Agency until January 31, 2021, and then according to three different hypothetical transmission scenarios.

We assume that 90% of all individuals older than 19 years will take the vaccine when offered, and a that 95% out of this group will be 100% protected by the vaccine and that 5% will have no protection (i.e., an all-or-nothing model with respect to a 95% vaccine efficacy). We do not consider vaccination of individuals younger than 20 years. People who recover from natural infection are assumed to be 100% protected. We assume no waning immunity. We do not consider changes in disease severity or reductions in vaccine efficacy associated with new strains.

### Scenario description

The model is used to simulate a number of different plausible scenarios varying four different factors:

I. Transmission scenario: The scenarios follow respectively three different trajectories (see Figure 1). These trajectories can be thought of as being affected by the new virus strain taking over, the level of prevention, or seasonal effects They are not affected by additional immunity from vaccination and should hence not be interpreted as *R*_*t*_. However, the realized *R*_*t*_ which arises from the scenario, and which is represented in our computations, takes immunity into account (see Figures S.8e to S.31e in S.M.). Transmission scenario 1 (T1) assumes no change from February 1, Transmission scenario 2 (T2) assumes a linear increase from 1 on February 1 up until April 30 when it has increased 30% (perhaps caused by invading virus strains), followed by a decrease (perhaps caused by seasonality), and Transmission scenario 3 (T3) equals T2 but with no decline from April 30.

**Figure 1.**
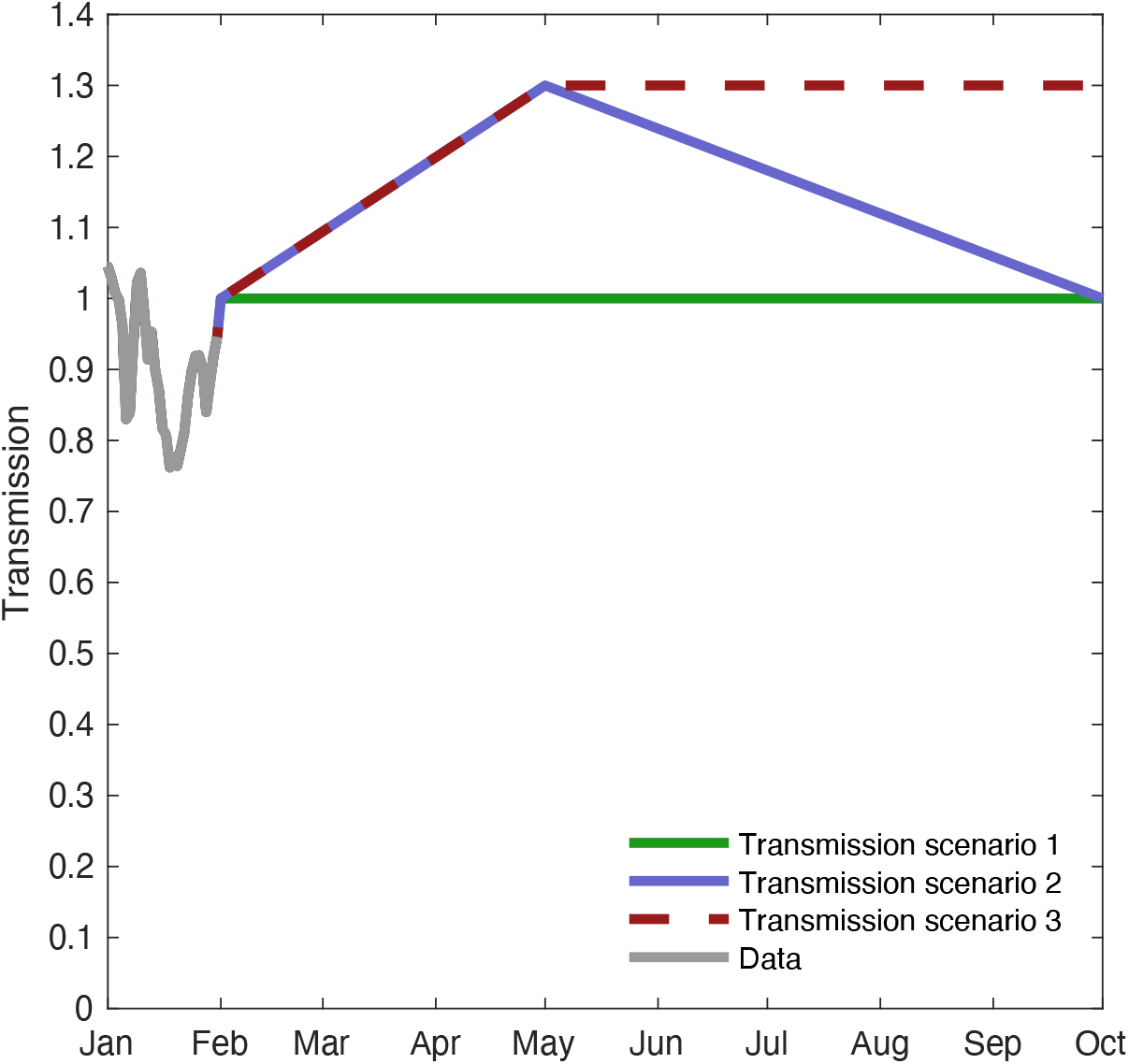
Model scenarios for Transmission rate. The grey values are estimated *R*_*t*_ values for Sweden (7). From the first of February we hypothesized three different scenarios for the transmission rate; Transmission scenario 1 (T1), Transmission scenario 2 (T2) and Transmission scenario 3 (T3). T1 assumes the transmission rate remains constant at 1 from first of February. Scenario T2 and T3 assume increased transmission rate up until end of April, either from relaxed compliance to recommendations and restrictions and/or increased dominance of new virus strains with higher transmissibility. Scenario T2 then assumes a reduced transmission from May due to seasonality, which T3 does not. The transmission scenario curves differ from *R*_*t*_ in that immunity from vaccination and disease transmission is not taken into account. The realized *R*_*t*_, influenced by the respective transmission scenarios, however, take this effect from immunity into account (see Figures S.8e to S.31e in S.M.).
II. Delay or no delay of vaccine deliveries: The anticipated increase of vaccine deliveries in beginning of April is delayed by one month or not (see Figure 2).

**Figure 2.**
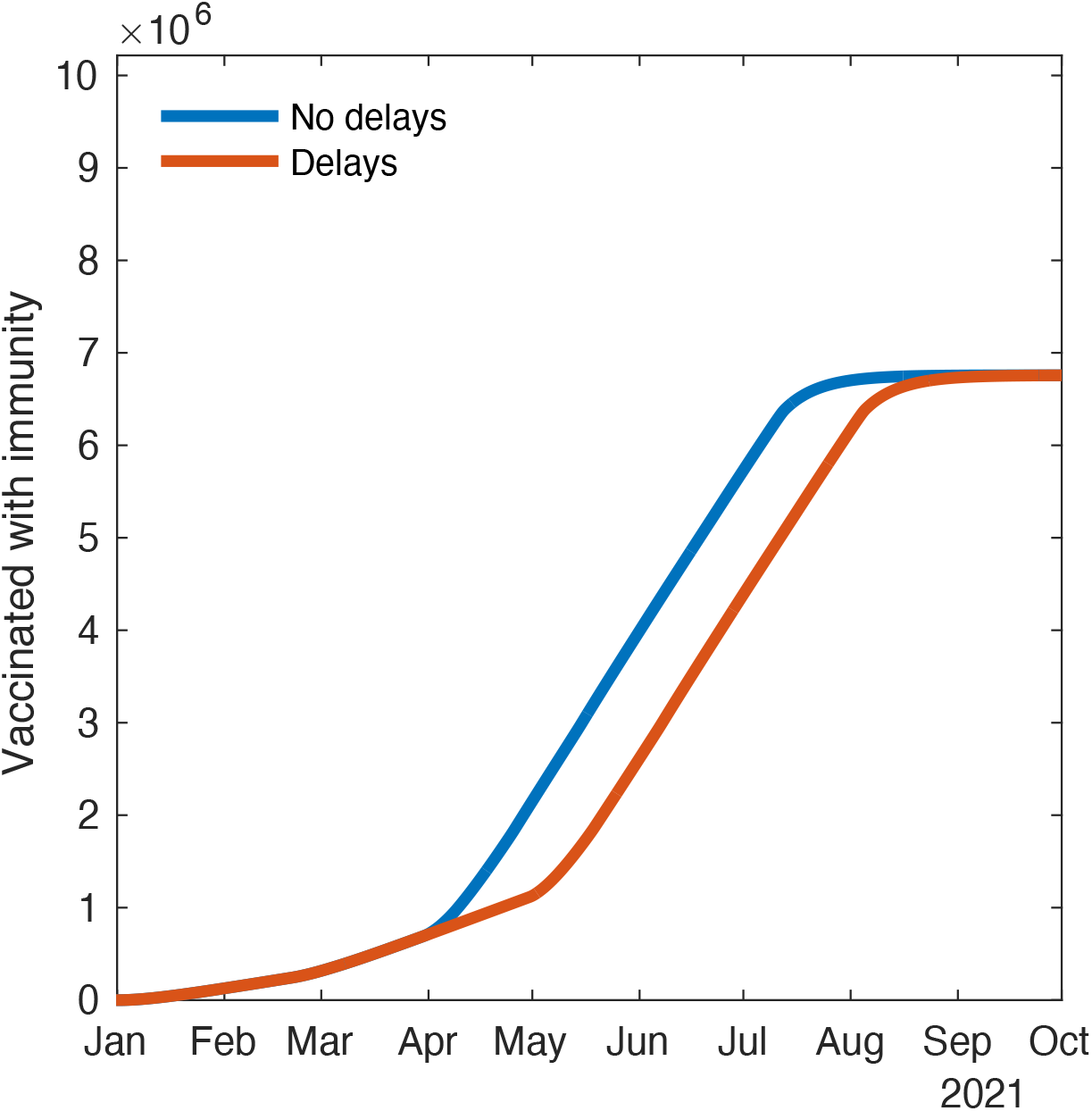
Vaccination delivery and uptake scenario for Sweden. The planned (blue) and the delayed (red) vaccination delivery scenario is, respectively, ramping up from May 1 or April 1. Here, vaccine is offered also to antibody positive individuals. Vaccine delivery in January to February follows approximately observed levels in Sweden.
III. Postponement or no postponement of vaccination of antibody positive individuals.
IV. Ascending or Descending age order of vaccination. In both scenarios 70+ are vaccinated first and those under 20 are not vaccinated at all. The Descending vaccination order then vaccinates 60-69, followed by 50-59 and so on, whereas as Ascending order vaccinates 20-29, followed by 30-39 and so on. A motivation for the Descending order strategy is to prioritize with respect to risk of serious illness, whereas a motivation for the Ascending order strategy is to indirectly protect those at a higher risk by reducing transmission as quickly as possible (10).

The first factor (Transmission scenarios) has three levels whereas the other three factors all have two levels, resulting in a total of 24 different scenario combinations (see Table S6 in section 2.3 in S.M.).

The focus of the analysis lies on learning how the different scenarios affect disease outcomes and how they can inform the development of an optimal strategy. For each scenario we analyse the progress of the epidemic up until October 1 with respect to four different outcomes: infections, regular health care, critical care, and fatalities. The parameters for in the analysis are informed by Swedish data, but the aim is not to give the best possible fit or forecast of the Swedish COVID-19 development.

## Results

Figure 3 shows the model results on the number of individuals in critical care over time, where we have varied one of the four factors (I-IV) at a time, where the other factors are kept at the following state: Transmission scenario 2, No vaccine delay, Vaccinating also antibody positive, and Descending age order vaccination.

**Figure 3.**
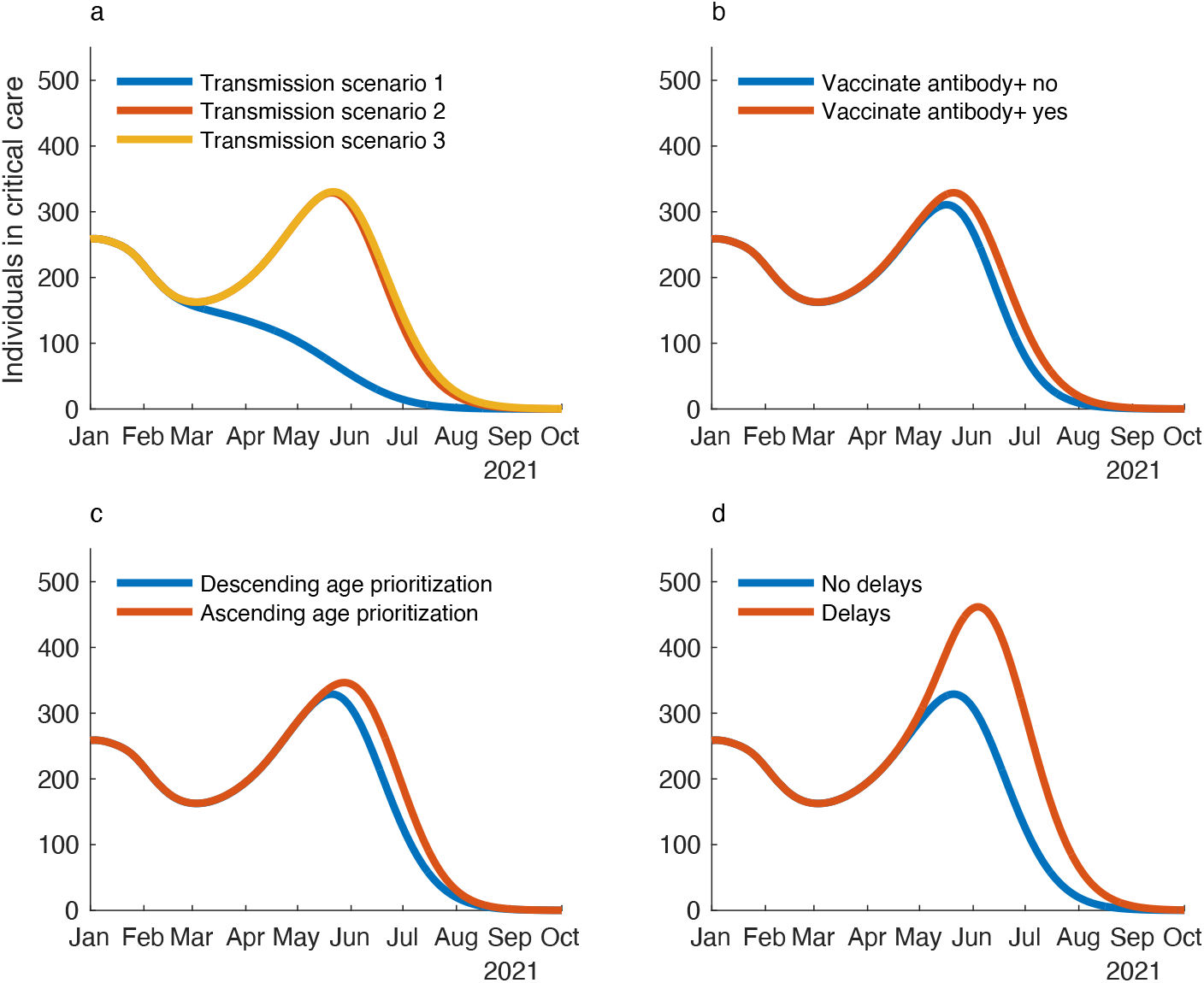
Patients in critical care over time. Factors not varied are kept at: Transmission scenario 2, No vaccine delay, Vaccinating also antibody positive, and Descending age order vaccination. a) Different transmission scenarios (T1 vs. T2 vs. T3). b) Postponement or no postponement of vaccination of antibody positive individuals. c) Ascending or Descending age order of vaccination. d) Delay or no delay of vaccine deliveries.

The factor with the largest impact on the number of individuals in critical care is the Transmission scenario (I). More specifically, if T1 happens, or either of T2 or T3. As a consequence, it is of utmost importance to reduce transmission in March and April. In contrast, the difference between T2 and T3 is very minor indicating that reducing transmission later is less significant for the critical care need since by then all individuals above 50 will have been vaccinated (given a descending age order vaccination). Of the remaining three factors (II, III, IV), the vaccine deliveries (II) are most important. Efficient deliveries (as well as quickly vaccinating once vaccines arrive) is hence also crucial. When it comes to prioritizing whom to vaccinate first (III, IV), the simulations suggest that it is best to vaccinate in descending age order, and to recommend individuals having natural antibodies to vaccinate only when susceptible individuals have been vaccinated.

In S.M. we show the corresponding results for infections, regular healthcare and fatalities (Figures S1 to S3), which show similar patterns but with two exceptions. First, fatalities have a much lower increase in all scenarios (due to vaccination of risk groups and elderly starting already in January, but see also Discussion). Second, when comparing Ascending and Descending age order of vaccination, the outcomes for regular healthcare, c ritical care and fatalities are lower (i.e., better) for a Descending age order of vaccination, whereas incidence is lower (i.e., better) for an Ascending age order of vaccination. As a consequence, vaccination in ascending order does reduce transmission more, but not enough to compensate for the large healthcare load as an effect of that individuals in ages 50+ are being vaccinated only later.

Figure 4 shows the relative reduction between March 1 and October 1 2021, for all four disease-burden outcomes, and all possible combination of scenarios (see S.M. section 1.5 for methodology). The most important factor in keeping health damage to a minimum is hence to keep transmission low. T1 rather than T3 reduces the number of individuals needing critical care by 60-73%. Second most important is timely vaccine deliveries and expeditious vaccination management upon delivery. By avoiding a one-month delay of vaccine deliveries, the number of patients needing critical care is reduced by 10-32%. Vaccinating in a descending age-order, instead of in an ascending age-order, reduces the number of patients needing critical care by 2-11%. Postponing vaccination of individuals with natural immunity has bigger impact than the vaccination order, and it number of patients needing critical care by 2-14%. The relative reduction effect from vaccinating in descending order is *negative* for number of cases; as mentioned above, this actually increases transmission (but decreases the remaining three health burden outcomes). See also Table S6 in S.M. section 2.3.

**Figure 4.**
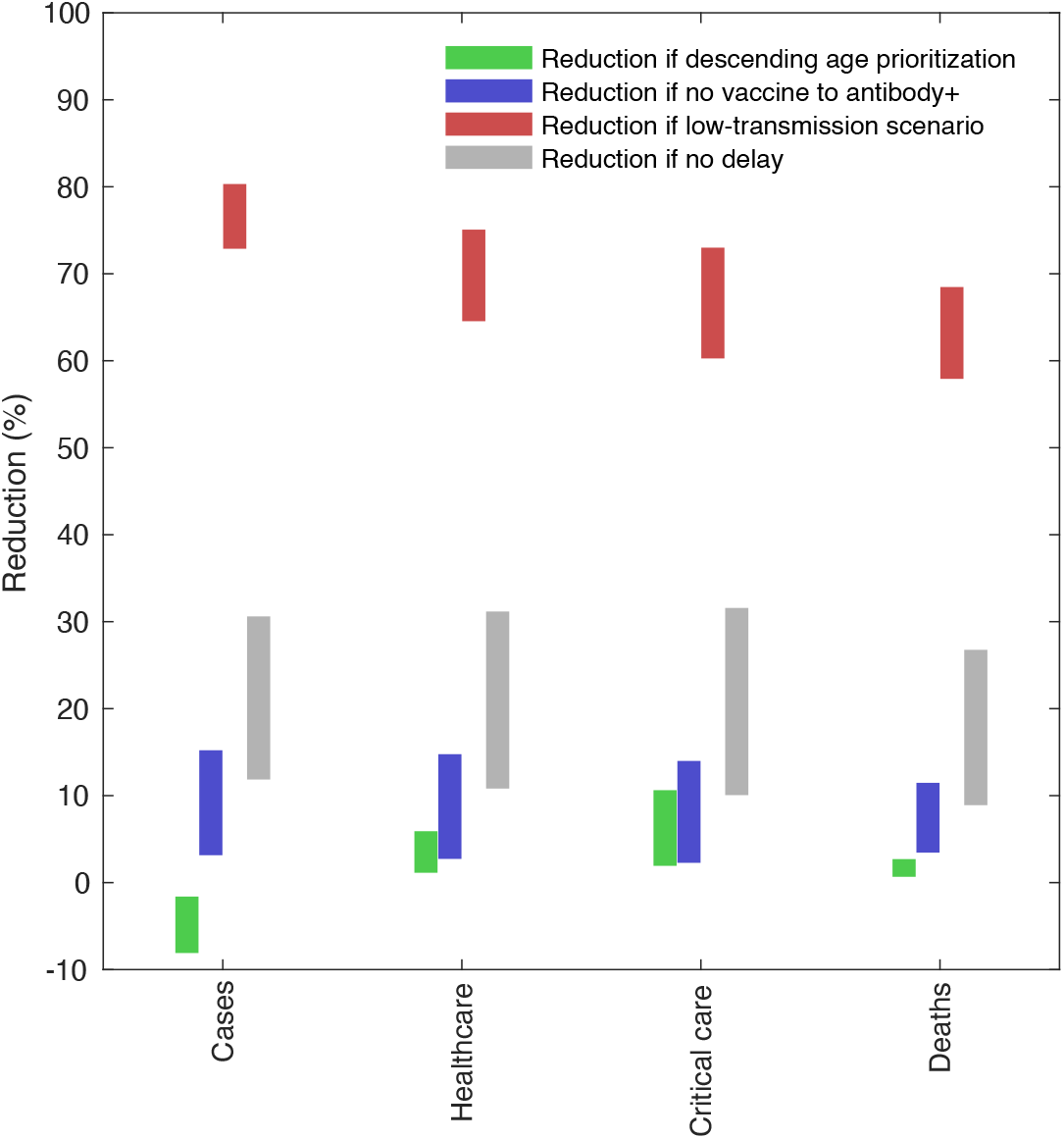
Relative reduction (%) for cumulative numbers of cases, person-days of healthcare, person-days of critical care, and deaths, for the age-order prioritization (red); delaying vaccination of people with antibodies (dark yellow); transmission scenario (beige); no delay in vaccination (grey).

In Figure 4 we also see that all ranges of relative effects exclude the null effect; meaning that the value zero is not contained in any interval. This result suggest that our results are quite robust to the factors studied, and thus possibly also to other model modifications which would imply that our results carry over to models containing more realistic features and also when adapted to other regions/countries not too different from Sweden.

In the supplement we also show the age distribution of the health care demands and deaths. When the numbers decrease as vaccination progresses, the cases in health care, critical care and deaths are shifting to overall lower ages compared to the pre-vaccination period (Figures S5 to S7 in S.M. section 2.2).

## Discussion

The results of our analysis suggest that keeping transmission rates low and administrating vaccinations without delays is the dominant factor for reducing the disease burden of COVID-19 infections. Our results also suggest that it is most important, whenever possible, to down-prioritize vaccinations among people with natural immunity. Vaccination prioritization with respect to age (after that 70+ age individuals have been vaccinated) is less important, yet vaccinating in a descending age order was shown to be slightly more effective than vaccinating in an ascending age order. Whereas the results of this study are likely generally applicable to a range of countries, it should foremost be seen as a case study for Sweden as our analysis was informed by Swedish data.

Clearly, our model is a rough simplification of the actual COVID-19 pandemic. Still, the model considers differences in contact patterns, severity risks and vaccination priorities between age groups, and contain hospital and intensive care and death or recovery from different health care units estimated from data. All potentially important features are not captured in the model. The model does not account for spatial heterogeneity, nor does it explain the high transmission among frail people above the age of 80+, for example in care homes. One explanation could be that the contact rates among this demographic group is larger than contacts otherwise in the same age group, and that also contact rates with some lower age-groups is increased due to frequent home service. This is not well captured in our study. As a consequence, the model underestimated deaths in Sweden, but only until this age group is fully protected by vaccines. Since our study objective was to evaluate the impact of vaccination strategies in ages below 70 years of age, after everyone at age 70 years and above had been vaccinated, we consider this mismatch to be of reduced importance to the study findings. We assume a mean of ten days from vaccination to full immunity and do not explicitly model two vaccine doses (1,2) – relating to the fact that the time spent in different states are expressed by constant exit rates leading to exponential durations that may not be realistic. With respect to transmission dynamics we do not consider age-dependent susceptibility or infectiousness (yet age-dependent contact rates). Further, we do not consider partial or waning of immunity. Naturally infected individuals are thus assumed 100% protected (11). We note that the latter has implications for the size of the effect of the severity-reduction (i.e., relative reduction) from delaying vaccinations for antibody-positive individuals as such natural protection may last for a more limited time. We do not capture higher severity and case fatality associated with new strains, as reported for B1.1.7 (4). Our modelling could thus be improved in several aspects to correspond even better to the real situation.

The main purpose of our analysis, however, was not to fit our model to data in the best possible way, but instead to study and learn about what we think are the most central qualitative properties of the future progress of the epidemic under a set of different relevant scenarios. Whereas we believe that our qualitative conclusions are primarily applicable in Sweden, they are likely applicable also to other countries with similar characteristics for COVID-19. We think that future studies could benefit from extending mainly in the directions of: spatial heterogeneity; age-structured degrees of social distancing; and, waning immunity, where the latter would be relevant when consider slightly larger time-scales. Hopefully, this current study can still contribute to better understanding how to plan and act during the vaccination phase in order to best reduce the potential disease-burden in this high-pace race against the pandemic.

## Data Availability

All data in the paper are either from open sources, from scientific literature or otherwise explained in the paper.

## References

1. Knoll MD, Wonodi C. Oxford–AstraZeneca COVID-19 vaccine efficacy. The Lancet, 2021, 397.10269: 72-74.

2. Polack FP, et al. Safety and efficacy of the BNT162b2 mRNA Covid-19 vaccine. New England Journal of Medicine, 2020, 383.27: 2603-2615.

3. Davies, Nicholas G., et al. Estimated transmissibility and impact of SARS-CoV-2 lineage B.1.1.7 in England. Science, 2021, 10.1126/science.abg3055

4. Bager P et al. Increased risk of hospitalisation associated with infection with SARS-CoV-2 lineage B.1.1.7 in Denmark. https://ssrn.com/abstract=3792894

5. Sjödin H, et al. COVID-19 healthcare demand and mortality in Sweden in response to non-pharmaceutical mitigation and suppression scenarios. International Journal of Epidemiology, 2020, 49.5: 1443–1453.

6. Prem K, Cook AR, Jit M Projecting social contact matrices in 152 countries using contact surveys and demographic data. PLoS Comput Biol, 2017, 13(9): e1005697. https://doi.org/10.1371/journal.pcbi.1005697

7. Folkhälsomyndigheten, 18 February, 2021: https://www.folkhalsomyndigheten.se/smittskydd-beredskap/utbrott/aktuella-utbrott/covid-19/statistik-och-analyser/

8. Svenska intensivvårdsregistret, SIR, 18 February, 2021: https://www.icuregswe.org/data--resultat/covid-19-i-svensk-intensivvard/

9. Sun K, et al. Transmission heterogeneities, kinetics, and controllability of SARS-CoV-2. Science, 2021, 371.6526.

10. Matrajt L, Eaton J, Leung T, Brown ER. Vaccine optimization for COVID-19: who to vaccinate first?, MedRxiv, 2020 https://www.medrxiv.org/content/10.1101/2020.08.14.20175257v3

11. Bubar, KM, et al. Model-informed COVID-19 vaccine prioritization strategies by age and serostatus. Science, 2021, 371.6532: 916-921.

