## Supplementary Material for "Evaluating and optimizing COVID-19 vaccination policies: a case study of Sweden"

### 1 Methods

#### 1.1 The age-structured SEIR epidemic model

We developed an extended age-structured SEIR epidemic model defined by a set of 162 (9 age-groups times 18 epidemiological compartments) differential equations (Eq. S.1). Figure S1 privides a schematic view of the compartmental model represented as a flow-graph for a single age-group (any of the respective 9). The only non-linear terms in the ODE system is the infection rate for each age group (and vaccine allocation policies that are affected by natural immunity and the number of individuals in the preceeding vaccination cohorts). The infection rate equals the fraction of susceptible individuals in the specific age group multiplied by a weighted sum of the fractions of infectious individuals in the other age groups, where the weights reflect both the size of these age groups but also the mixing rate between the two age groups in question – a quantity which is country specific and taken from a social study (1).

A short summary overview. After an individual becomes infected, the individual is either destined to take the track for hospital-care or the home-care track. The probability for taking the hospital care track is age-structured and derived from data. Infected individuals are at first on average in a latent phase for three days, followed by a transmission phase of on average four days after which they remain medically infectious yet going into self-quarantine that is assumed to be 100% efficient. After some additional days they go into hospital care if on the hospital care track, or if not they recover and become immune. Individuals who enter hospitals have again an age-structured risk of being moved to an intensive care unit from which they recover via further hospital care or die with age-structured probabilities. Individuals below the age of 70 die only while being patients at an intensive care unit, whereas as older people also is exposed to a risk of dying in ordinary health care and also in home care. The age-structured rates and average durations are derived from data is given in Tables S2 and S3 and otherwise provided in the text. Individuals that recover are assumed completely immune for the rest of the study period.

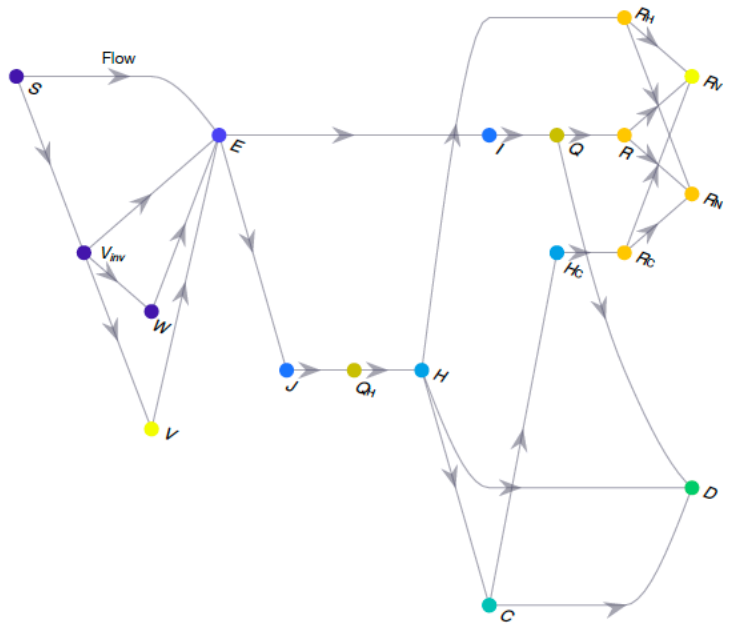


Figure S 1 Flow diagram of the extended SEIR model corresponding to Eq. S.1.
See Table S1.1 for state-variable notations.

The vaccination process was studied under 24 various scenarios which is given by permuations of factors I, II, III and IV as described in the main text. In the model, individuals that are offered a vaccine either accept the offer (adhere) or not, we assume 90% adherence. Those that accept the offer are vaccinated, and 95% out of them are assumed to become completely immune whereas the remaining 5% receive no effect from the vaccine (so called all-or-nothing model). Also this immunity is assumed to remain for the rest of the study period. The immunity is assumed to take full effect 10 days after the vaccination shot on average. We do not include two doses in our model.

More specifically, the extended SEIR model that we developed can be written as a system of ordinary equations, namely as

$$
\begin{aligned}
\frac{dS_a}{dt} &= -f_a(t)\, S_a - v_a(\breve{a})\, S_a \\
\frac{dE_a}{dt} &= (S_a + V_{inv} + W_a)\, f_a(t) - \frac{E_a}{T_L} \\
\frac{dI_a}{dt} &= \frac{(1 - p_H - p_C)\, E_a}{T_L} - \frac{I_a}{T_{trans}} \\
\frac{dJ_a}{dt} &= \frac{(p_H + p_C)\, E_a}{T_L} - \frac{J_a}{T_{trans}} \\
\frac{dQ_a}{dt} &= \frac{I_a}{T_{trans}} - \frac{Q_a}{T_Q} \\
\frac{dQ_{H\,a}}{dt} &= \frac{J_a}{T_{trans}} - \frac{Q_{H_a}}{T_{Q_H}} \\
\frac{dH_a}{dt} &= \frac{Q_{H_a}}{T_{Q_H}} - \frac{H_a}{T_H} \\
\frac{dC_a}{dt} &= \frac{\frac{p_C}{p_C + p_H}\, H_a}{T_H} - \frac{C_a}{T_C} \\
\frac{dH_{C\,a}}{dt} &= \frac{(1 - \mu_C)}{T_C}\, C_a - \frac{H_{C_a}}{T_{H_C}} \\
\frac{dD_a}{dt} &= \frac{\mu\, Q_a}{T_Q} + \frac{\mu_H \left(1 - \frac{p_C}{p_C + p_H}\right) H_a}{T_H} + \frac{\mu_C\, C_a}{T_C} \\
\frac{dR_{H\,a}}{dt} &= \frac{\left(1 - \frac{p_C}{p_C + p_H}\right)(1 - \mu_H)}{T_H} H_a - v_a(\breve{a})\, R_{H_a} \\
\frac{dR_{C\,a}}{dt} &= \frac{(1 - \mu_C) C_a}{T_C} - v_a(\breve{a})\, R_{C_a} \\
\frac{dR_a}{dt} &= \frac{(1 - \mu_O)}{T_Q}\, Q_a - v_a(\breve{a})\, R_a \\
\frac{dR_{N\,a}}{dt} &= v_a(\breve{a})\, (1 - \rho)\left(R_a + R_{H_a} + R_{C_a}\right) \\
\frac{dV_{inv\,a}}{dt} &= v_a(\breve{a})\, S_a - \left(f_a(t) + \frac{1}{T_{V_{inv}}}\right) V_{inv_a} \\
\frac{dV_a}{dt} &= \frac{\alpha\, e}{T_{V_{inv}}}\, V_{inv_a} \\
\frac{dW_a}{dt} &= \frac{(1 - \alpha\, e)}{T_{V_{inv}}}\, V_{inv_a} - f_a(t)\, W_a \\
\frac{dR_{V\,a}}{dt} &= v_a(\breve{a})\, \rho \left(R_a + R_{H_a} + R_{C_a}\right).
\end{aligned}
\qquad \text{(S.1)}
$$

The “force of infection” $f_a(t)$ is equal to the per-capita rate at which individuals in age-group $a$ contract the disease, and can be written as

$$f_a(t) = \frac{\tau(t)}{N_a} \sum_{i=1}^{9} c_{ia}\, \phi_i \qquad \text{(S.2)}$$

where $\phi_i$ denotes the number of infectious individuals in age-group $i$, $N_a$ the number of individuals in age-group $a$, $c_{ia}$ the per-capita rate at which individuals in age-group $i$ get into contact with individuals in age-group $a$ (see Table S2), and $\tau(t)$ is a scaling factor quantifying the transmission probability (this scaling factor also takes into account seasonal effects as well as effects of preventive measures reducing contact rates). Beside $\tau(t)$, the r.h.s. hence equals the number of infectious individuals in age-group $i$ that a single individual in age-group $a$ is in contact with per day. The time-dependent parameter

can be seen as a product between the probability that transmission occurs at a contact and a scaling-factor to the per-capita contact rate $c_{ia}$. The effective reproduction number

$$R(t) = \tau(t)\, \lambda \qquad \text{(S.3)}$$

where $\lambda$ is the largest eigenvalue of the matrix with any $ia$-th element equal to

$$c_{ia}\, (1 - \epsilon_a) T_{trans} \qquad \text{(S.4)}$$

where $\epsilon_a$ denotes the fraction of the population in age-group $a$ that is immune, $T_{trans} = 4$ the average time during which any infected individual transmits disease. From Eq. S.3 we can accordingly solve for $\tau(t)$ and obtain a value of $\tau(t)$ that corresponds to $R(t)$. One can thus easily force $\tau(t)$ either to correspond to a hypothetical value of $R(t)$ at time $t$, or to an empirically estimated value of $R(t)$. We used empirically estimated daily values of $R(t)$ for November 30 2020 < t < February 1 2021, and thereafter altered between three hypothetical transmission scenarios, see Figure 1 in the main text.

The per-capita vaccination rate of age-group $a$ can be written as

$$v_a(\breve{a}) = \omega(a, \xi) \left(1 - \frac{\omega(\breve{a}, \xi)\, \psi(\breve{a})}{\theta}\right) \qquad \text{(S.5)}$$

where $\theta$ denotes the vaccination-rate capacity and $\psi(\breve{a})$ the number of yet unvaccinated individuals in the vaccination cohort $\breve{a}$ preceeding cohort $a$. When there are no individuals left to vaccinate in the preceeding vaccination cohort $\breve{a}$, then $v_a(\breve{a})$ is equal to the per-capita rate

$$\omega(a, \xi) = \frac{\theta}{\psi(a) + \xi\, \theta} \qquad \text{(S.6)}$$

where the parameter $\xi$ governs the relation between the number of individuals left to vaccinate, the vaccination-rate capacity, and the per-capita vaccination rate. As an example, $v_a(\breve{a}) = \omega(a, \xi)\, /2$ when $\psi(\breve{a}) = \xi\theta$. We chose $\xi = 1/5$. The number of not yet vaccinated individuals equals

$$\psi(a) = S_a + R_a + R_{H_a} + R_{C_a} \qquad \text{(S.7)}$$

for the scenarios where antibody-positive individuals reveive vaccinations, and

$$\psi(a) = S_a \qquad \text{(S.8)}$$

for the scenarios where antibody-positive individuals do not reveive vaccinations (see Table S6). The factor $v_a(\breve{a})$ thus introduce non-linearity to the terms where it occurs in the ode-system (Eq. S.1).

| **Table S1. Notation and description of state variables in Eq. S.1.** | |
|---|---|
| **State variable** | **Description** |
| $S_a$ | Susceptible |
| $E_a$ | Exposed |
| $I_a$ | Infected (not going to hospital) |
| $J_a$ | Infected (going into hospital) |
| $H_a$ | In healthcare |
| $C_a$ | In critical care |
| $H_{C_a}$ | In recovery ward after critical care. |
| $D_a$ | Dead |
| $Q_a$ | In quarantine (at "home") |
| $R_{H_a}$ | Recovered after healthcare (antibody+) |
| $R_{C_a}$ | Recovered after critical care (antibody+) |
| $R_a$ | Recovered at "home" (antibody+) |
| $V_{inv_a}$ | Susceptibles invited to take vaccine |
| $V_a$ | Vaccinated susceptibles that developed immunity |
| $W_a$ | Susceptibles with no immune response or declined vaccination |
| $R_{N_a}$ | Non-vaccinated antibody+ individuals |
| $R_{V_a}$ | Vaccinated antibody+ individuals |
| $Q_{H_a}$ | In quarantine before hospital admission |

#### 1.2 Parameters

The contact matrix was taken from (1) and reflects contact patterns for Sweden in year 2017. Each element $c_{ia}$ in the matrix reflects the average number of close contacts an individual in age group $i$ has with individuals in age group $a$ per day. For example, $c_{42} = 1.31$ implies an individual of age 31-40 on average have 1.31 close contacts with individuals of age 10-19 per day.

**Table S2. Contact matrix for 10 year age groups in Sweden taken from (1). The row reflects the age of the individual and the column the type of contacts considered, and elements of the table reflect average number of close contacts per day. See text above for more information.**

| Age groups | 0-9 | 10-19 | 20-29 | 30-39 | 40-49 | 50-59 | 60-69 | 70-79 | 80+ |
|---|---|---|---|---|---|---|---|---|---|
| 1: 0-10 | 5,17 | 0,90 | 0,80 | 1,97 | 0,92 | 0,43 | 0,34 | 0,12 | 0,1 |
| 2: 11-20 | 0,99 | 9,23 | 1,38 | 1,29 | 1,82 | 0,56 | 0,18 | 0,09 | 0,1 |
| 3: 21-30 | 0,58 | 1,87 | 5,79 | 2,89 | 2,32 | 1,44 | 0,21 | 0,09 | 0,1 |
| 4: 31-40 | 1,62 | 1,31 | 2,53 | 5,05 | 3,16 | 1,56 | 0,42 | 0,12 | 0,1 |
| 5: 41-50 | 0,90 | 2,18 | 2,12 | 3,25 | 4,35 | 1,77 | 0,35 | 0,15 | 0,1 |
| 6: 51-60 | 0,96 | 1,79 | 2,16 | 2,58 | 3,06 | 3,05 | 0,64 | 0,18 | 0,1 |
| 7: 61-70 | 0,79 | 0,70 | 0,90 | 1,43 | 1,16 | 1,17 | 2,22 | 0,51 | 0,1 |
| 8: 71-80 | 0,46 | 0,71 | 0,38 | 0,69 | 0,95 | 0,73 | 1,16 | 1,21 | 0,3 |
| 9: 80+ | 0,23 | 0,22 | 0,26 | 0,25 | 0,25 | 0,25 | 0,64 | 1,11 | 0,6 |

Table S3 provides the complete set of parameter values used in the ode-system Eq S.1. The infectious period and latent period was taken from published data (5). The time to quarantine was taken to be two days after symtoms onset, thus 4 days in total from onset of infectiousness (5). This accounted for that a smaller proportion of asymptomatics may not quarantine at all. The pre-hospitalization time, i.e. the time from desease onset to hospitalization, has been estimated to 7 days (2). The average time in hospital has likewise been estimated to 7 days (2). Whereas the average time spent in critical-care was substantially longer initially in the pandemic with a median around 20 day (6), it has recently been estimated to around 9 days (7,8). The average time in hospital recovering from critical-care has not been well documentet. We therefore set the average time in post critical-care recovery only to one day to downplay its influence due to the uncertainties in its duration. The immunization latency time was set lower than expected to avoid large exponentially distributed that are likely not realistic. The time in quarantine is emergent from the time to quarantine. The death risk among out-of-hospital patients was unknown and calibrated to data. The deaths risk among hospitalized patients by age group was derived from a combination of (2,3) where the total critical care deaths by age group was substracted from all deaths in in-patient care. The deaths risk among critical care patients was derived directly from data (2,3).

**Table S3. Notation, description, estimate and reference for model parameter values.**

| | | Age group | | | | | | | | |
|---|---|---|---|---|---|---|---|---|---|---|
| Notation | Description | 0-9 | 10-19 | 20-29 | 30-39 | 40-49 | 50-59 | 60-69 | 70-79 | 80+ |
| $T_L$ | latent period | 3,00 | 3,00 | 3,00 | 3,00 | 3,00 | 3,00 | 3,00 | 3,00 | 3,00 |
| $T_{INF}$ | Infectious period | 9,00 | 9,00 | 9,00 | 9,00 | 9,00 | 9,00 | 9,00 | 9,00 | 9,00 |
| $T_{trans}$ | Time to quarantine | 4,00 | 4,00 | 4,00 | 4,00 | 4,00 | 4,00 | 4,00 | 4,00 | 4,00 |
| $T_{preH}$ | Pre-hospitalization time | 7,00 | 7,00 | 7,00 | 7,00 | 7,00 | 7,00 | 7,00 | 7,00 | 7,00 |
| $T_H$ | Hospitalization period | 7,00 | 7,00 | 7,00 | 7,00 | 7,00 | 7,00 | 7,00 | 7,00 | 7,00 |
| $T_C$ | Critical care period | 9,00 | 9,00 | 9,00 | 9,00 | 9,00 | 9,00 | 9,00 | 9,00 | 9,00 |
| $T_{H_C}$ | Post critical care hospital period | 1,00 | 1,00 | 1,00 | 1,00 | 1,00 | 1,00 | 1,00 | 1,00 | 1,00 |
| $T_{inv}$ | Immunization latency time | 10,00 | 10,00 | 10,00 | 10,00 | 10,00 | 10,00 | 10,00 | 10,00 | 10,00 |
| $T_Q = T_{INF} - T_{trans}$ | Time in quarantine | 5.00 | 5.00 | 5.00 | 5.00 | 5.00 | 5.00 | 5.00 | 5.00 | 5.00 |
| $T_{Q_H} = T_{preH} - T_{trans}$ | Time in quarantine before hospitalization | 3.00 | 3.00 | 3.00 | 3.00 | 3.00 | 3.00 | 3.00 | 3.00 | 3.00 |
| $\mu$ | Fraction of non-hospitalized that dies | 0,00 | 0,00 | 0,00 | 0,00 | 0,00 | 0,00 | 0,00 | 0,08 | 0,20 |
| $\mu_H$ | Fraction of hospitalized that dies | 0,00 | 0,00 | 0,00 | 0,00 | 0,00 | 0,00 | 0,00 | 0,20 | 0,40 |
| $\mu_C$ | Fraction of inds. In critical care that dies | 0,35 | 0,10 | 0,10 | 0,15 | 0,15 | 0,22 | 0,46 | 0,49 | 0,52 |
| $p_H$ | Fraction of infected needing health care | 0,00 | 0,00 | 0,02 | 0,03 | 0,04 | 0,08 | 0,16 | 0,43 | 0,52 |
| $p_C$ | Fraction of infected needing critical care | 0,00 | 0,00 | 0,00 | 0,00 | 0,00 | 0,01 | 0,03 | 0,05 | 0,01 |
| $e$ | Vaccine efficacy | 0,95 | 0,95 | 0,95 | 0,95 | 0,95 | 0,95 | 0,95 | 0,95 | 0,95 |
| $\alpha$ | Maximum vaccination coverage | 0,00 | 0,00 | 0,90 | 0,90 | 0,90 | 0,90 | 0,90 | 0,90 | 0,90 |

The fraction of infecected requiring hospital care was derived from data on hospital admissions (a, b) divided by the an estimation the cumulative number of infected (Table S.4) and multiplied by a factor to account for underreporting. The overall under-reporting was assumed to equal 50% compared to the reported number of cases. This was assumed unevenly distributed among age groups. More specifically we assumed that the under-reporting in different age-groups were inverse proportional, i.e., $1 + 9\,k^{-2.46}$, to the age-group index $k = 1,2, \dots, 9$ (thus implying much more under-reporting among children and low in older age-groups). The vaccine efficacy was assumed to be 95% and the maximium vaccination coverage was assumed to be zero for individuals under age 20 (as they were not considered fro vaccination) and otherwise 90% motivated by a 90% vaccination adherence.

The inverse proportional underreporting was motivated by the very large case fatality rates observed in younger age groups, and the difference between the estimated infection fatality ratio and the case fatality ratio. The fraction of individuals requiring healthcare was set based on healthcare data from (2,3), and the fraction of individuals requiring critical care was based on (7,8).

| | **Table S4. Data and estimates of the number of cumulative confirmed cases, and the number of individuals with health care need or critical care need from COVID-19 by age groups in Sweden up to 17th January, 2021*** | | | | | | | | | |
|---|---|---|---|---|---|---|---|---|---|---|
| Notation | Description | Values | | | | | | | | |
| $a$ | Age group | 0-10 | 11-20 | 21-30 | 31-40 | 41-50 | 51-60 | 61-70 | 71-80 | 80+ |
| $\mathbb{C}$ | Confirmed cases | 6745 | 52557 | 93288 | 91576 | 94784 | 87617 | 46268 | 23114 | 27482 |
| $\mathcal{H}$ | Health care** | 205 | 374 | 1640 | 2122 | 3197 | 5088 | 5430 | 7046 | 9992 |
| $\mathcal{C}$ | Critical care | 17 | 31 | 136 | 176 | 442 | 1008 | 1306 | 1106 | 296 |
| $\mathcal{D}$ | Deaths | 7 | 3 | 15 | 32 | 79 | 259 | 709 | 2578 | 8644 |
| $\mathcal{I}$ | Infections *** | 67450 | 138525 | 149567 | 118800 | 111059 | 97224 | 49740 | 24363 | 28593 |
| $\mathcal{H}_f = \frac{\mathcal{H}}{\sum \mathcal{H}}$ | Normalized $\mathcal{H}$ | 0.0058 | 0.0107 | 0.0467 | 0.0605 | 0.0911 | 0.1450 | 0.1547 | 0.2008 | 0.2847 |
| $\mathcal{C}_f = \frac{\mathcal{C}}{\sum \mathcal{C}}$ | Normalized $\mathcal{C}$ | 0.0038 | 0.0069 | 0.0301 | 0.0390 | 0.0978 | 0.2231 | 0.2891 | 0.2448 | 0.0655 |
| $\mathcal{D}_f = \frac{\mathcal{D}}{\sum \mathcal{D}}$ | Normalized $\mathcal{D}$ | 0.0006 | 0.0002 | 0.0012 | 0.0026 | 0.0064 | 0.0210 | 0.0575 | 0.2092 | 0.7013 |
| $\mathcal{I}_f = \frac{\mathcal{I}}{\sum \mathcal{I}}$ | Normalized $\mathcal{I}$ | 0.0859 | 0.1764 | 0.1905 | 0.1513 | 0.1414 | 0.1238 | 0.0633 | 0.0310 | 0.0364 |

*Data was taken from open sources (2,3,4).
** Health care by ages groups was estimated from cumulative health care numbers by looking at in-patient data by age group from the National Board of Health and Welfare and substracting the total number of patients by age group of those being critical care patients.
*** The estimated total number of infections by the 17th of January, 2021, was derived from data on confirmed cases by January 17 2021, and up-scaled by a factor $1 + 9\,k^{-2.46}$ with the age-group index $k = 1,2, \dots, 9$.

We used time series data for the period 1 of December 2020 to 18th of February 2021 for validating model results on incidence, deaths, patients in critical care, and patients in health care. Data on incidence, deaths, patients in critical care available form was taken from (2), and data on patients in health care from (4).

#### 1.3 Initial conditions

The initial conditions were chosen to mimic the situation in Sweden as of December 1, 2020 to the best of our knowledge. Below we present and motivate our choices.

We constructed the initial condition

$$
\begin{aligned}
S &= N - (E + I + J + H + C + H_C + D + Q + R_H + R_C + R + V_{inv} + V + W + R_N \\
&\qquad + R_V + Q_H) \\
E &= (I + J)\frac{T_L}{T_{trans}} \\
I &= h\,\mathcal{H}_f\,\frac{T_{trans}}{T_H}\frac{(1 - p_H - p_C)}{(p_H + p_C)} \\
J &= h\,\mathcal{H}_f\,\frac{T_{trans}}{T_H} \\
Q &= I\,\frac{T_Q}{T_{trans}} \\
Q_H &= J\,\frac{T_{Q_H}}{T_{trans}} \\
H &= h\,\mathcal{H}_f \\
C &= h_c \mathcal{C}_f \\
H_C &= C\,(1 - \mu_C)\frac{T_{H_C}}{T_C} \\
D &= d\,\mathcal{D}_f \\
R_H &= r\,p_H\,(1 - \mu_H)\,\mathcal{I}_f \\
R_C &= r\,p_C\,(1 - \mu_C)\,\mathcal{I}_f \\
R &= r\,(1 - p_H - p_C)\,(1 - \mu)\mathcal{I}_f \\
R_N &= 0 \\
V_{inv} &= 0 \\
V &= 0 \\
W &= 0 \\
R_V &= 0
\end{aligned}
\tag{S.9}
$$

directly from or derived from data. All notation is explained in Table S3, and Table S4, except for the cumulative number of infections $r$ December 1, the number of patiants in healthcare $h$ December 1, the number of patients in critical care $h_c$ December 1, and the number of fatalities $d$ up until December 1. All state variables are assumed to be age-structured vectors.The corresponding numerical initial comditions are given in Table S5.

**Table S5. Initial conditions for the different compartment at the 1th of December, 2020**

| | 0-10 | 11-20 | 21-30 | 31-40 | 41-50 | 51-60 | 61-70 | 71-80 | 80+ |
|---|---|---|---|---|---|---|---|---|---|
| $S_a$ | 1208566 | 1042569 | 1135916 | 1129038 | 1084004 | 1086842 | 1001357 | 896922 | 437432 |
| $E_a$ | 184 | 1436 | 2549 | 2502 | 2590 | 2394 | 1264 | 632 | 751 |
| $I_a$ | 459 | 3578 | 6267 | 6119 | 6279 | 5684 | 2849 | 1148 | 1208 |
| $J_a$ | 0 | 2 | 76 | 99 | 155 | 249 | 271 | 338 | 451 |
| $H_a$ | 12 | 22 | 96 | 125 | 188 | 299 | 319 | 414 | 587 |
| $C_a$ | 1 | 2 | 7 | 9 | 23 | 54 | 69 | 59 | 16 |
| $H_{C_a}$ | 0 | 0 | 1 | 1 | 3 | 12 | 32 | 29 | 8 |
| $D_a$ | 4 | 2 | 9 | 19 | 47 | 153 | 420 | 1527 | 5121 |
| $Q_a$ | 459 | 3578 | 6267 | 6119 | 6279 | 5684 | 2849 | 1148 | 1208 |
| $R_{H_a}$ | 430 | 785 | 3444 | 4457 | 6714 | 10685 | 11404 | 14798 | 20984 |
| $R_{C_a}$ | 24 | 45 | 195 | 253 | 635 | 1448 | 1876 | 1589 | 425 |
| $R_a$ | 13141 | 102398 | 181755 | 178419 | 184669 | 170706 | 90145 | 45033 | 53544 |
| $V_{inv_a}$ | 0 | 0 | 0 | 0 | 0 | 0 | 0 | 0 | 0 |
| $V_a$ | 0 | 0 | 0 | 0 | 0 | 0 | 0 | 0 | 0 |
| $W_a$ | 0 | 0 | 0 | 0 | 0 | 0 | 0 | 0 | 0 |
| $R_{N_a}$ | 0 | 0 | 0 | 0 | 0 | 0 | 0 | 0 | 0 |
| $R_{V_a}$ | 0 | 0 | 0 | 0 | 0 | 0 | 0 | 0 | 0 |
| $Q_{H_a}$ | 0 | 1 | 33 | 42 | 66 | 107 | 116 | 145 | 193 |
| $N_a$ | 1223275 | 1154417 | 1336614 | 1327201 | 1291653 | 1284315 | 1112971 | 963781 | 521929 |

$N_a$ = Total population
$R_{H_a} + R_{C_a} + R_a$ is equal to 0.11 $N_a$, i.e., 11% of the population in each age-group $a$.

#### 1.4 Numerical integration

We solved equation system S.1 numerically using MATLAB R2020b. We wrote the MATLAB-code, and it will be made available on request and at a public GitHub repository upon publication.

#### 1.5 Relative reduction

We refer to the relative positive effect that each respective factor (I, II, II, IV) has on each respective cumulative severity measure (infections, healthcare, critical care, deaths) as the relative reduction. We defined the relative reduction $r_{ij}$ for scenario factor $i$ and for scenario $j$ as

$$r_{ij} = \frac{X_{ijb}(t_1, t_2) - X_{ija}(t_1, t_2)}{X_{ijb}(t_1, t_2)}, \tag{S.10}$$

where $X_{ijk}(t_1, t_2)$ denote cumulative number of a severity measure from time $t_1$ to time $t_2$, and for factor variant $k = a, b$ where $a$ and $b$ are the two contrast alternatives in scenario factor $i$. The contrast alternatives were accordingly chosen so that a positive $r_{ij}$ refers to a reduction and a negative $r_{ij}$ refers to the opposite, i.e., an increase. More specifically, with respect to the cumulative severity measures: For infections, $X_{ijk}(t_1, t_2)$ refers to the cumulative number of infections. For healthcare, $X_{ijk}(t_1, t_2)$ refers to the number of person-days of healthcare (i.e., the sum of persons in healthcare per day over all days between $t_1$and $t_2$). For critical care, $X_{ijk}(t_1, t_2)$ refers to the number of person-days of critical care (i.e., the sum of persons in critical care per day over all days between $t_1$ and $t_2$). For deaths, $X_{ijk}(t_1, t_2)$ refers to the number of fatalities.

We can easily for any scenario factor $i$ compare the relative reduction between all possible combinations by varing the remaining scenario factors. Figure 4 in the main text show the result of all possible combinations. See additionally Table S6 for all 24 scenarios considered and the respective outcome of severity measures.

### 2 Results

#### 2.1 Time dynamics of disease severity outcomes under different scenarios

The time dynamics for critical care was presented in the main text (Figure 3). Below we present the corresponding figures for incidence (Figure S2), health care (excluding critical care) (Figure S3) and deaths (Figure S4).

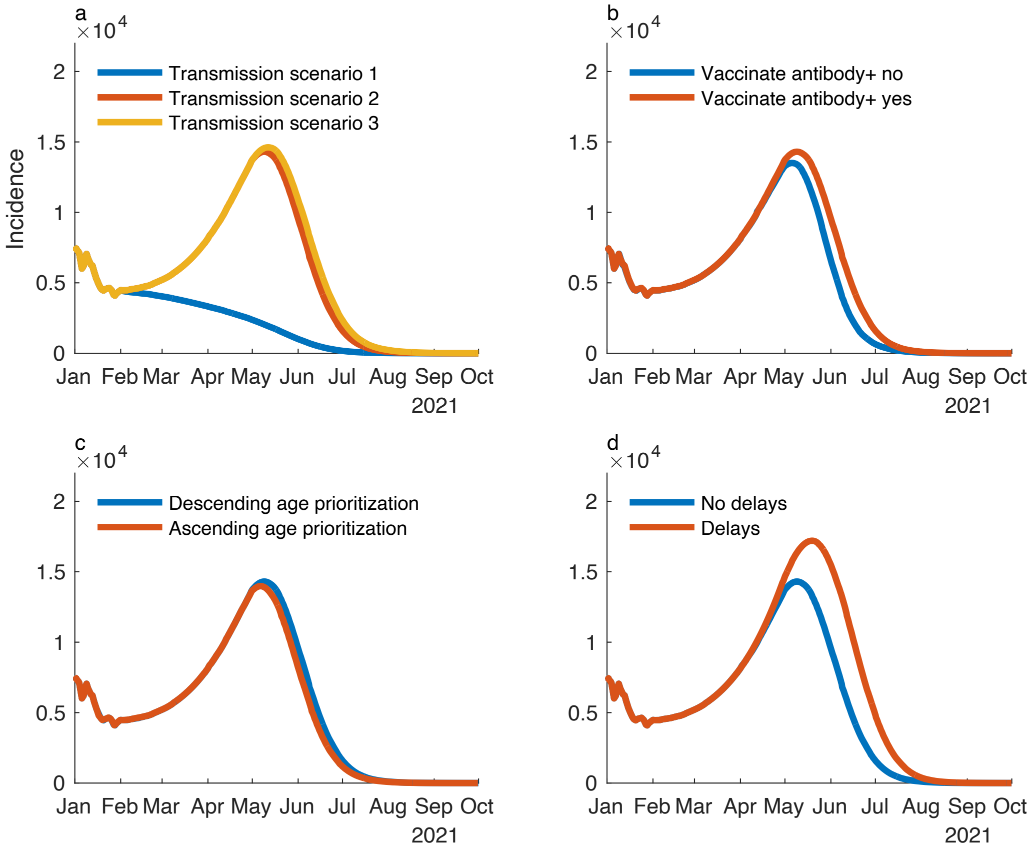


Figure S 2. Incidence over time, varying one factor in each sub-figure keeping other factors at: T2, Vaccinating antibody+, Descending vaccination order and No vaccination delay.

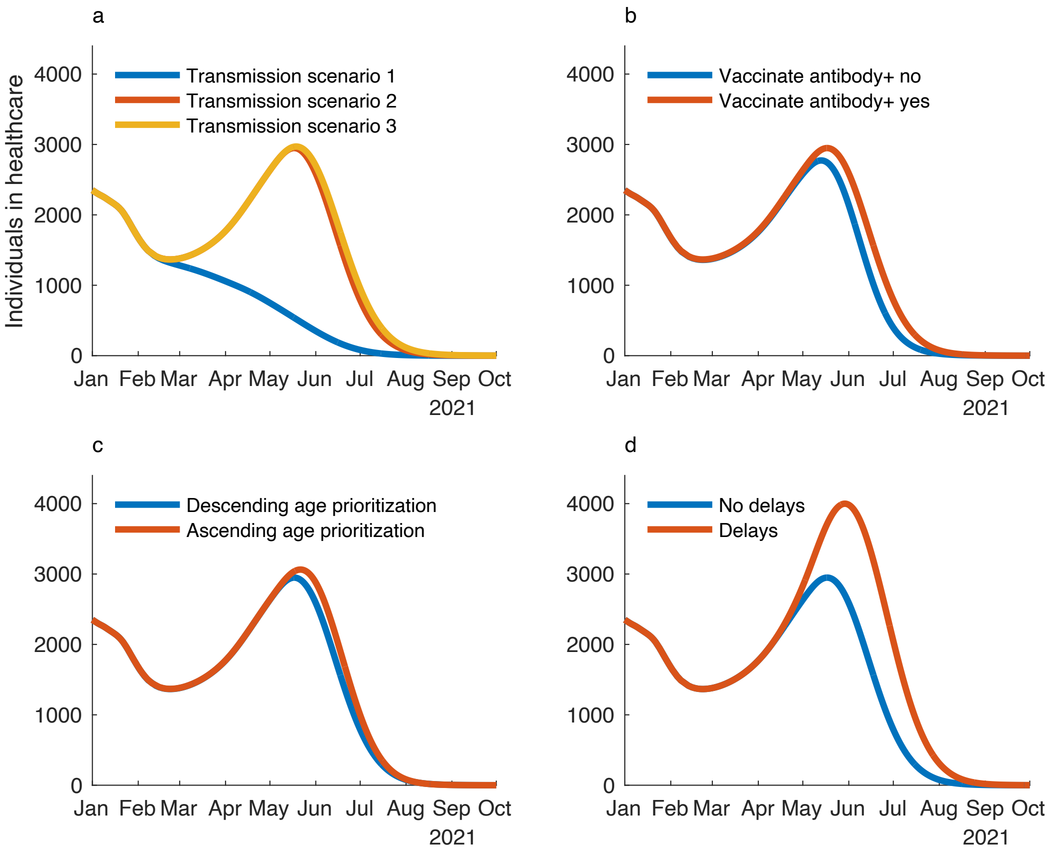


Figure S 3. Individuals in health care over time, varying one factor in each sub-figure keeping other factors at: T2, Vaccinating antibody+, Descending vaccination order and No vaccination delay.

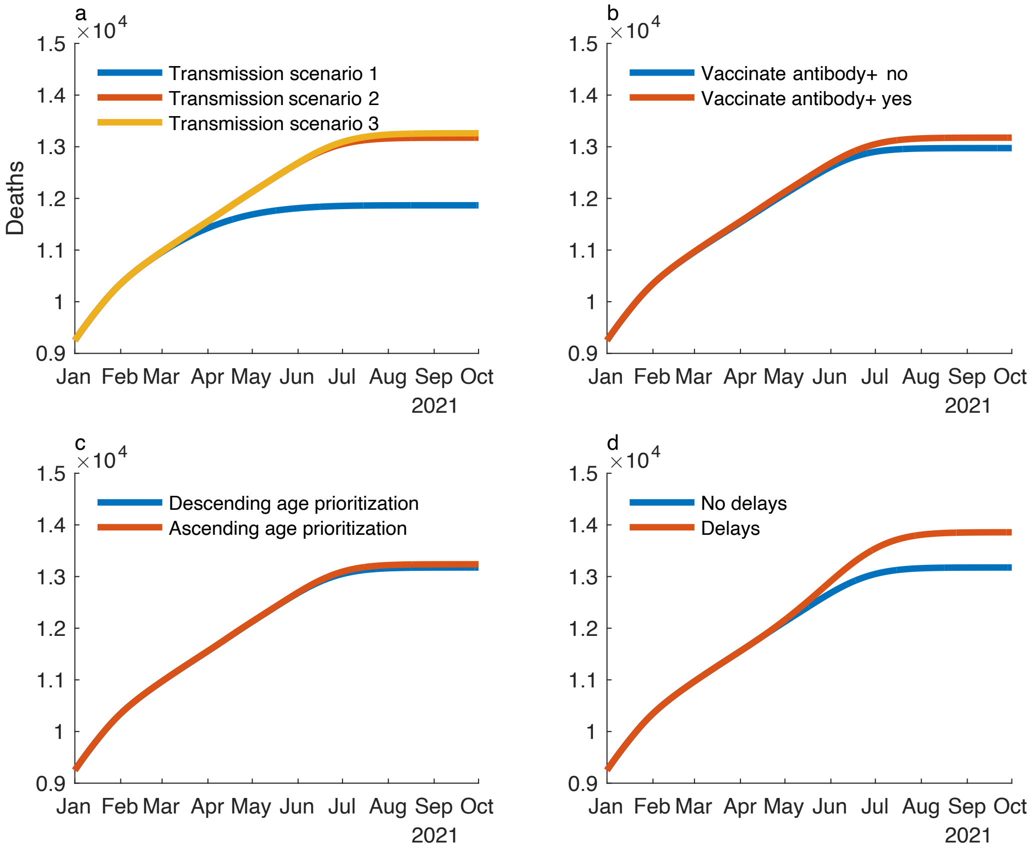


Figure S 4. Cumulative number of deaths over time, varying one factor in each sub-figure keeping other factors at: T2, Vaccinating antibody+, Descending vaccination order and No vaccination delay.

#### 2.2 Age distribution of individuals in healthcare, critical care, and deaths

The figures below report the age distribution of patient-days in healthcare and critical care, and the number of individuals who have died, during the period March 1 to October 1 2021, under the 24 different scenarios. The average number of days spent in health care equals 7 days, and in critical care 9 days, so the corresponding cumulative number of patients are obtained by dividing by 7 and 9 respectively.

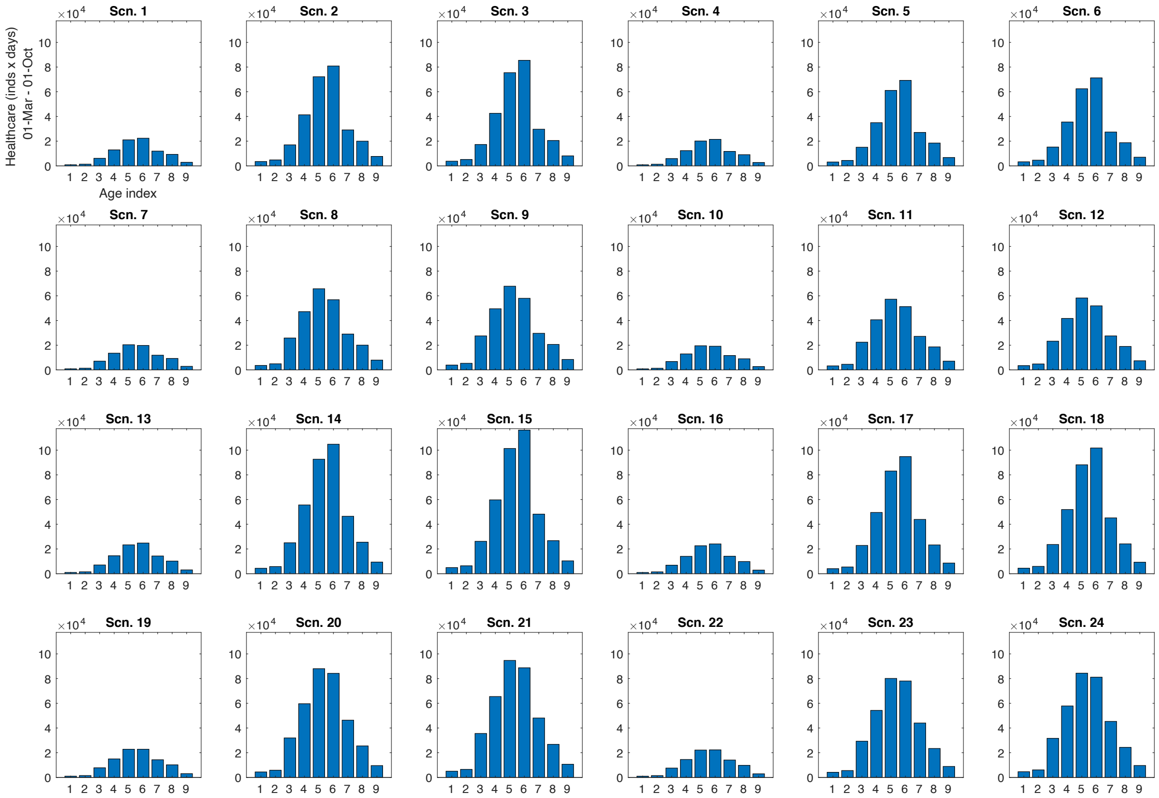


Figure S 5 Age distribution of patient-days spent in health care between March 1 and October 1, under the 24 different scenarios. The first group correspond to 0-9 years old, group 2 to 10-19, and so on.

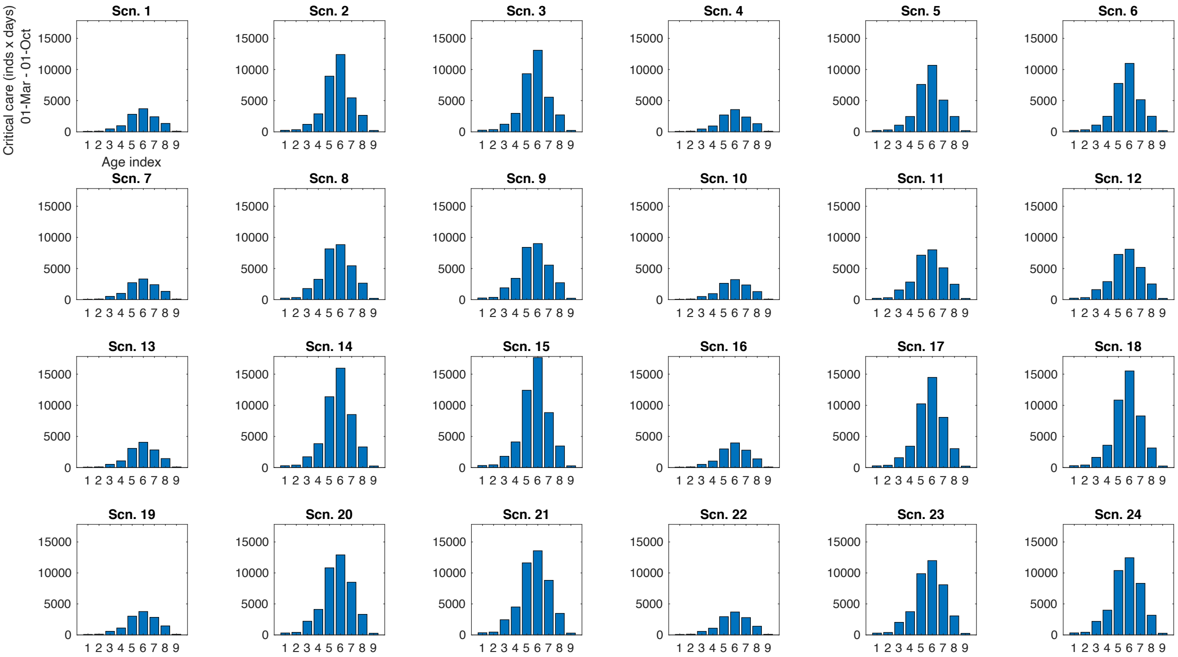


Figure S 6 Age distribution of number of patient-days spent in critical care between March 1 and October 1, under the 24 different scenarios.

.

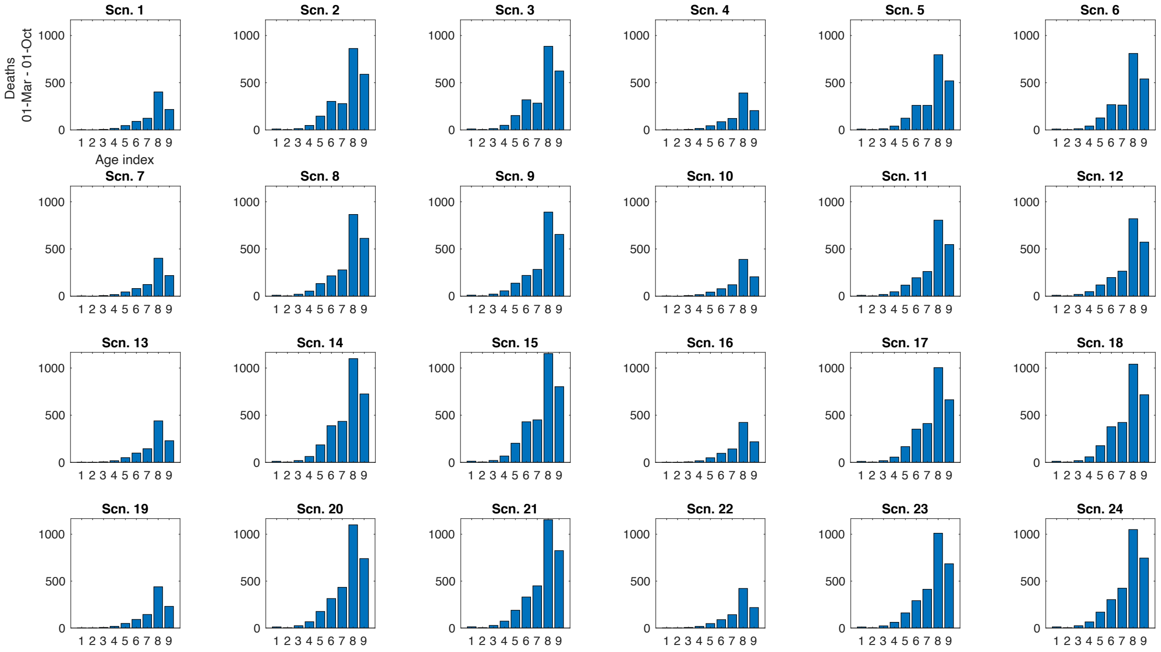


Figure S 7 Age distribution of number of deaths between March 1 and October 1, under the 24 different scenarios.

#### 2.3 Table for Cumulative outcomes of the 24 scenarios

The table describe the four outcomes: cumulative number of infected persons, total number of patient-days of health care (HC-days), patient-days in critical care (ICU-days), and the number of deaths (Deaths) associated with the 24 scenarios and counting from March 1 to October 1, 2021. The other columns reflect: scenario number (Sc-Id), the vaccination order (Order), vaccinating directly or postponing individuals with natural antibody response (VaccAB), transmission scenario (Tr-scn), delay in vaccine delivery and uptake (Delay).

**Table S6. Cumulative number of infected, patient-days in health care, patient-days in critical care and deaths, between March 1 and October 1, for the 24 scenarios.**

| ID | Order | VaccAB+ | Tr-scn | Delay | Infected | HC-days | ICU-days | Deaths |
|---|---|---|---|---|---|---|---|---|
| 1 | Asc | Directly | Tr-scn 1 | No | 263684 | 88974 | 11993 | 904 |
| 2 | Asc | Directly | Tr-scn 2 | No | 1034948 | 276440 | 34328 | 2253 |
| 3 | Asc | Directly | Tr-scn 3 | No | 1091312 | 288015 | 35725 | 2342 |
| 4 | Asc | Postp | Tr-scn 1 | No | 250952 | 85568 | 11606 | 872 |
| 5 | Asc | Postp | Tr-scn 2 | No | 894268 | 240136 | 30090 | 2025 |
| 6 | Asc | Postp | Tr-scn 3 | No | 925268 | 245496 | 30728 | 2073 |
| 7 | Desc | Directly | Tr-scn 1 | No | 270197 | 87339 | 11633 | 896 |
| 8 | Desc | Directly | Tr-scn 2 | No | 1092832 | 261323 | 31007 | 2193 |
| 9 | Desc | Directly | Tr-scn 3 | No | 1161574 | 270990 | 31930 | 2278 |
| 10 | Desc | Postp | Tr-scn 1 | No | 257804 | 84210 | 11290 | 863 |
| 11 | Desc | Postp | Tr-scn 2 | No | 960336 | 232426 | 27921 | 2002 |
| 12 | Desc | Postp | Tr-scn 3 | No | 1000140 | 237500 | 28412 | 2053 |
| 13 | Asc | Directly | Tr-scn 1 | Yes | 301686 | 99765 | 13334 | 992 |
| 14 | Asc | Directly | Tr-scn 2 | Yes | 1370713 | 369516 | 45721 | 2934 |
| 15 | Asc | Directly | Tr-scn 3 | Yes | 1503792 | 400171 | 49383 | 3146 |
| 16 | Asc | Postp | Tr-scn 1 | Yes | 291675 | 96870 | 13005 | 958 |
| 17 | Asc | Postp | Tr-scn 2 | Yes | 1245690 | 335588 | 41765 | 2690 |
| 18 | Asc | Postp | Tr-scn 3 | Yes | 1333332 | 354392 | 44012 | 2830 |
| 19 | Desc | Directly | Tr-scn 1 | Yes | 306537 | 98455 | 13050 | 985 |
| 20 | Desc | Directly | Tr-scn 2 | Yes | 1408832 | 355568 | 42849 | 2874 |
| 21 | Desc | Directly | Tr-scn 3 | Yes | 1557145 | 381559 | 45522 | 3070 |
| 22 | Desc | Postp | Tr-scn 1 | Yes | 296878 | 95771 | 12753 | 949 |
| 23 | Desc | Postp | Tr-scn 2 | Yes | 1298627 | 327801 | 39732 | 2662 |
| 24 | Desc | Postp | Tr-scn 3 | Yes | 1401204 | 345046 | 41524 | 2801 |

#### 2.4 Time dynamics for the 24 scenarios; cases, care, ICU, deaths

Below the time dynamics of the model is presented for each of the 24 scenarios. We report the incidence (infections), the cumulative number of deaths, patients in healthcare and critical care, the transmission scenarios including $R_t$ and the vaccine uptake. The gray curves up to February 17 reflect the corresponding observed numbers (where incidence is obtained from reported cases by assuming age-specific under-reporting as decribed in Section S1.2). In general, the model-fit to data is satisfactory for health care and critical care, whereas the model clearly under-estimates deaths (the model is calibrated to incidence/ $R_t$-data). There are many possible explanations to this under-estimation: to low age-specific mortality risks, higher under-reporting in older age-groups, prevalence in December and January being particularly high in older age-groups, slower vaccine uptake, and in general the model being too simple. As a consequence, the number of deaths under the different scenarios are not reliable, but we believe the ratio when comparing different scenarios still are.

The two lower curve reflect the transmission scenarios (left) and the vaccine uptake (right) for the specific scenario. The transmission scenario is equal to the reported $R_t$-values up until February 18. After this date, the transmission scenario (blue) is as hypothesized. The lower red curve is the corresponding $R_t$-value which also takes both natural and vaccine-induced immunity into account. The vaccination curves report the number individuals who have been vaccinated and who are immune thereafter. It hence includes individuals who were vaccinated and who were antibody positive (from disease-exposure) prior to vaccination whereas it excludes vaccinated individuals which did not develop immunity (assumed to equal 5%). Finally, for the vaccination strategy where antibody positive individuals are vaccinated last, these individuals are not contained in the graph, explaining why these vaccination curve end at about 5.5 million as compared to 6.8 million. Remember that individuals below 20 are not vaccinated, some do not adhere and some do not develop immunity, explaining why the number does not come close the the overall population size of 10.3 million.

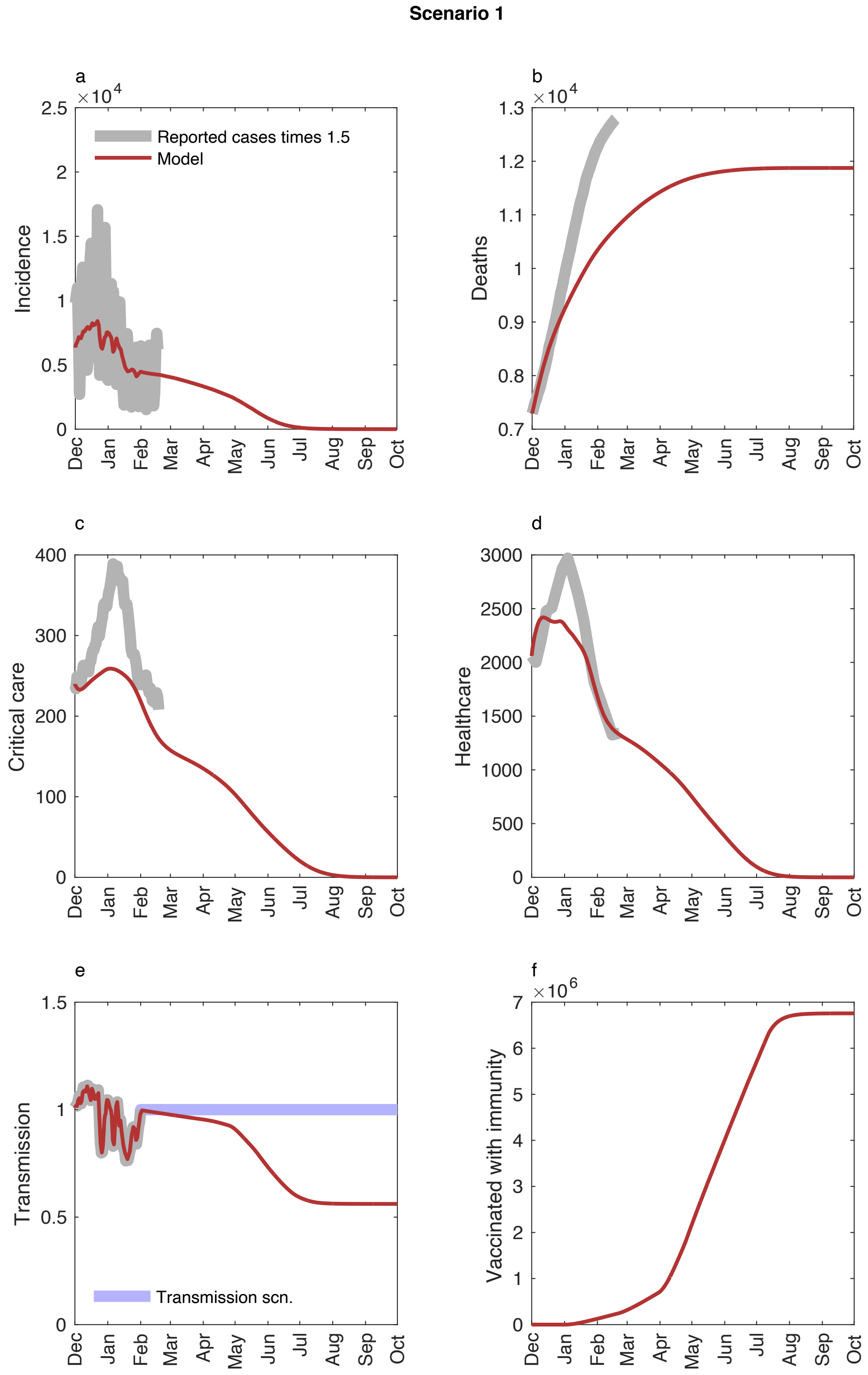


268
269 Figure S 8. Model dynamics for Model scenario 1. Upper left: number of infected, upper right: cumulative
270 deaths, mid left: patients in health care, mid right: patients in critical care, lower left: transmission scenario
271 (blue) and R(t) (red), lower right: number of vaccinated with immunity. See text for additional information.

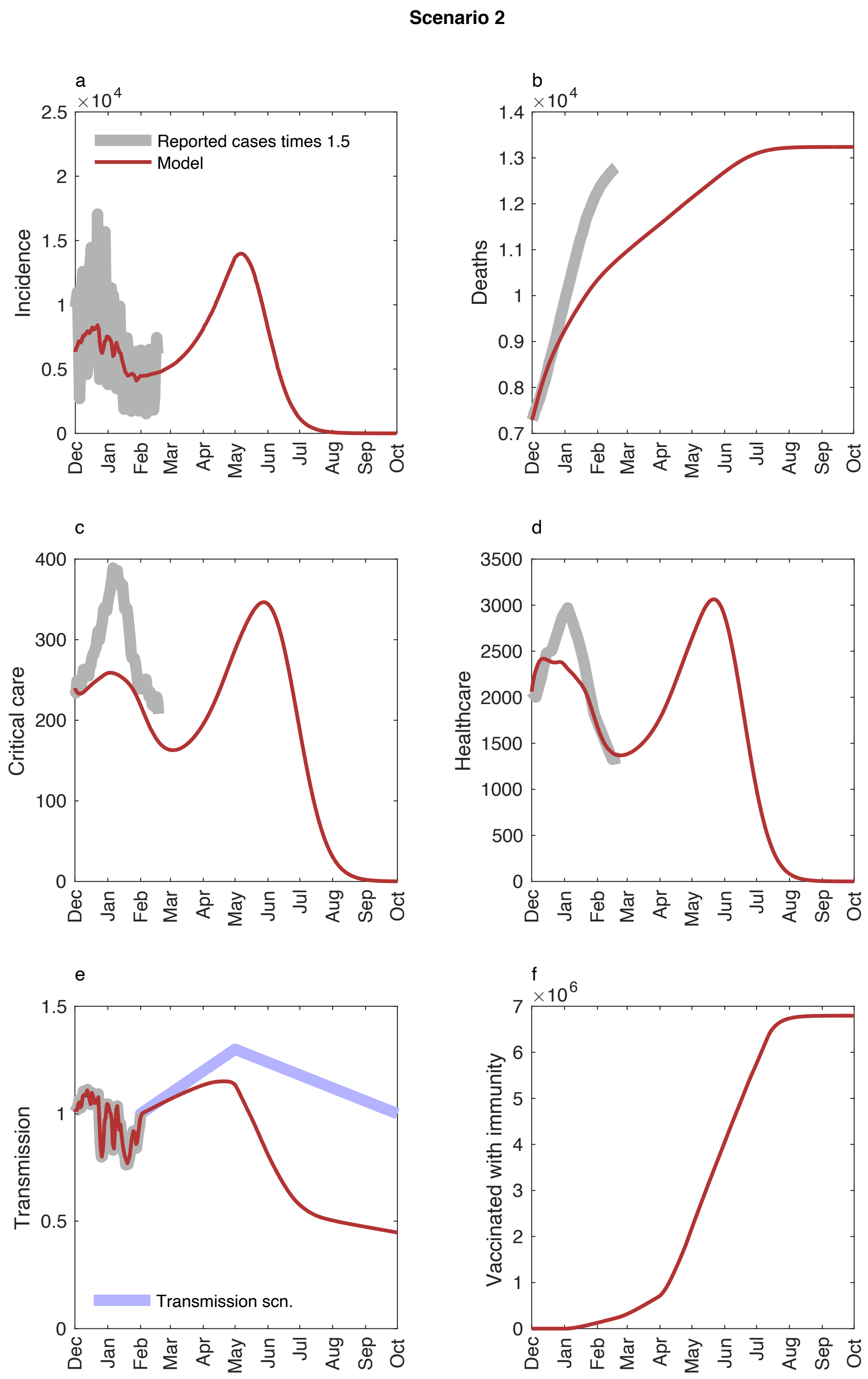


Figure S 9. Model dynamics for Model scenario 2. Upper left: number of infected, upper right: cumulative deaths, mid left: patients in health care, mid right: patients in critical care, lower left: transmission scenario (blue) and R(t) (red), lower right: number of vaccinated with immunity. See text for additional information.

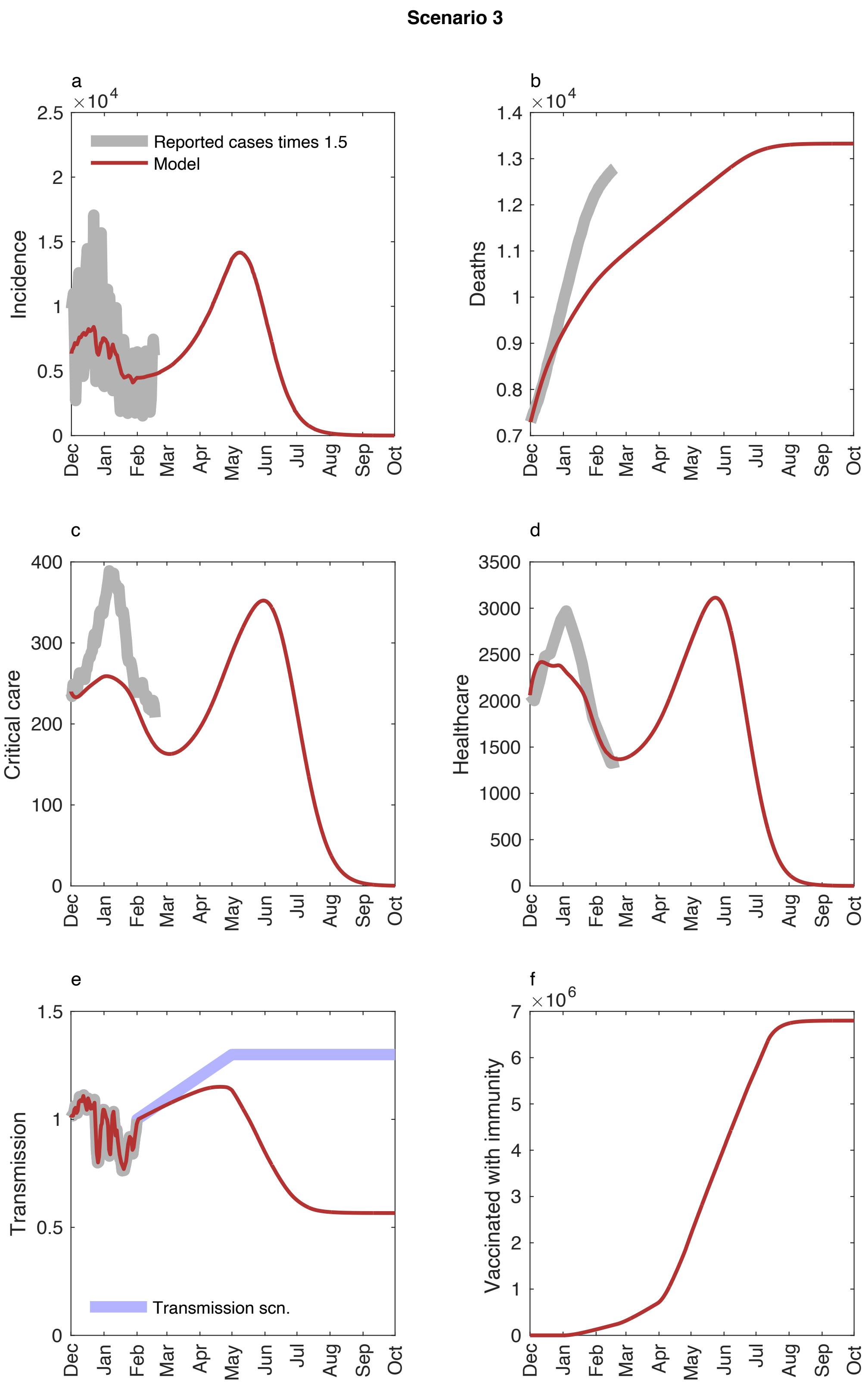


Figure S 10. Model dynamics for Model scenario 3. Upper left: number of infected, upper right: cumulative deaths, mid left: patients in health care, mid right: patients in critical care, lower left: transmission scenario (blue) and R(t) (red), lower right: number of vaccinated with immunity. See text for additional information.

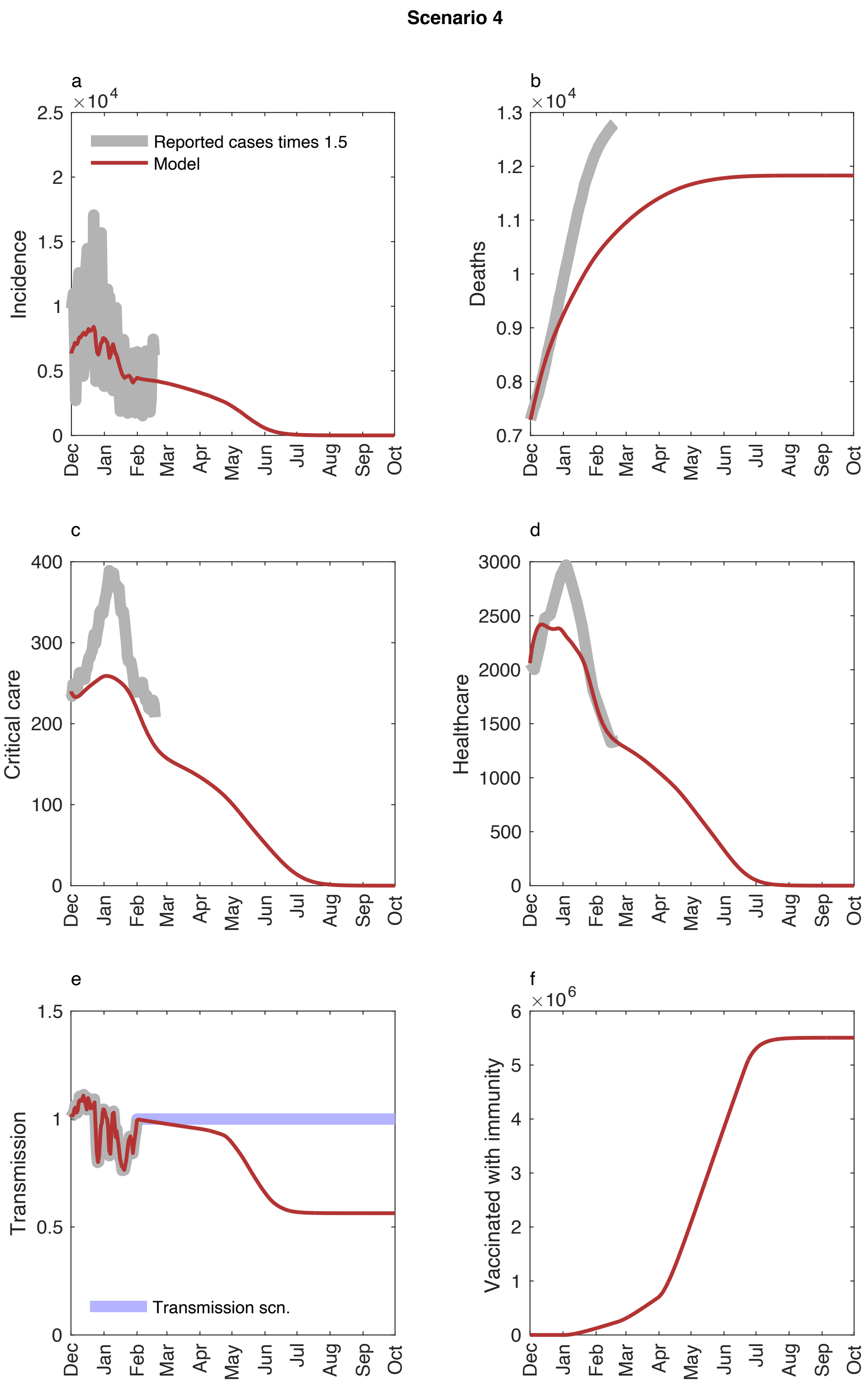


Figure S 11. Model dynamics for Model scenario 4. Upper left: number of infected, upper right: cumulative deaths, mid left: patients in health care, mid right: patients in critical care, lower left: transmission scenario (blue) and R(t) (red), lower right: number of vaccinated with immunity. See text for additional information.

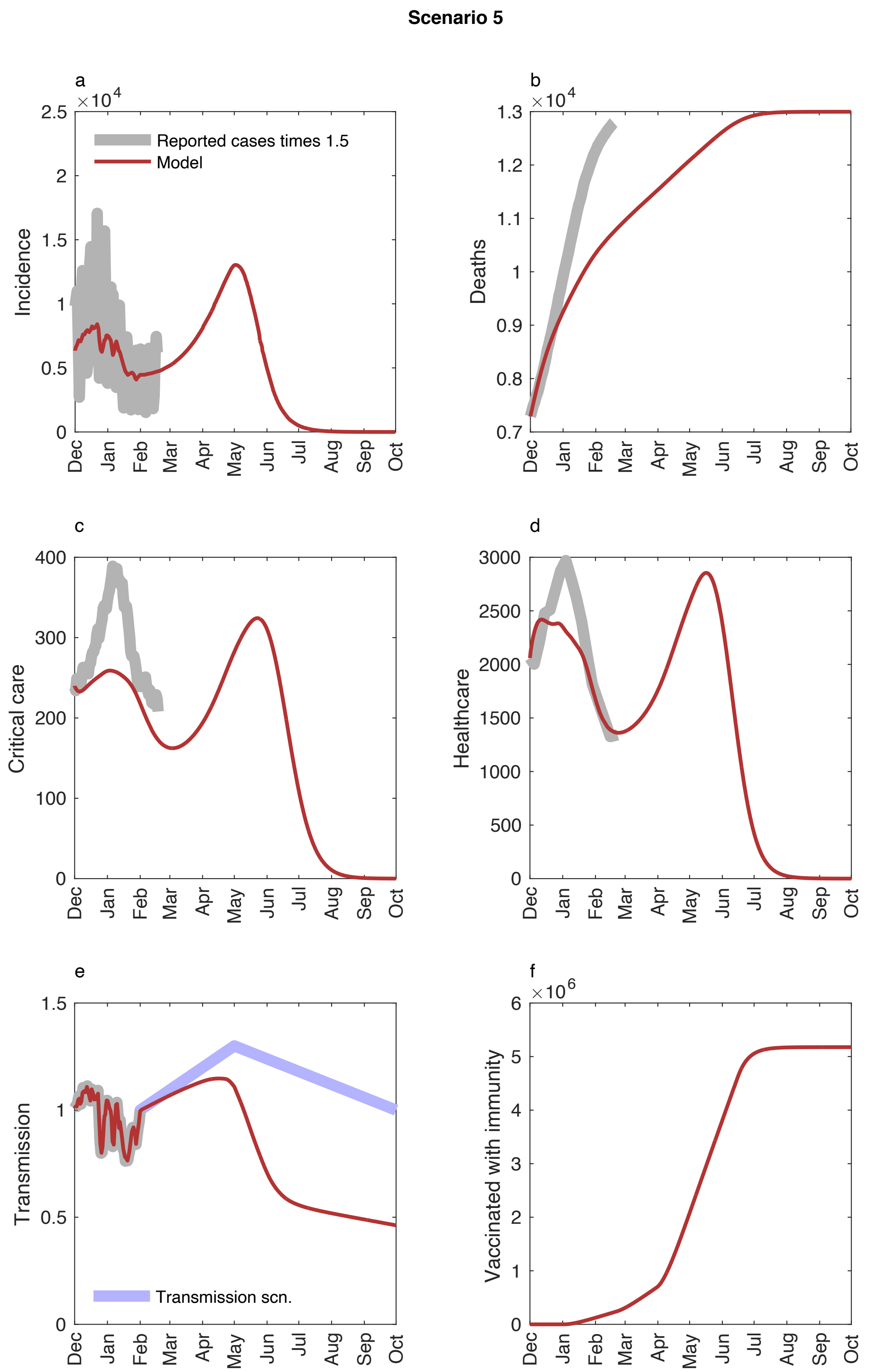


Figure S 12. Model dynamics for Model scenario 5. Upper left: number of infected, upper right: cumulative deaths, mid left: patients in health care, mid right: patients in critical care, lower left: transmission scenario (blue) and R(t) (red), lower right: number of vaccinated with immunity. See text for additional information.

Scenario 6

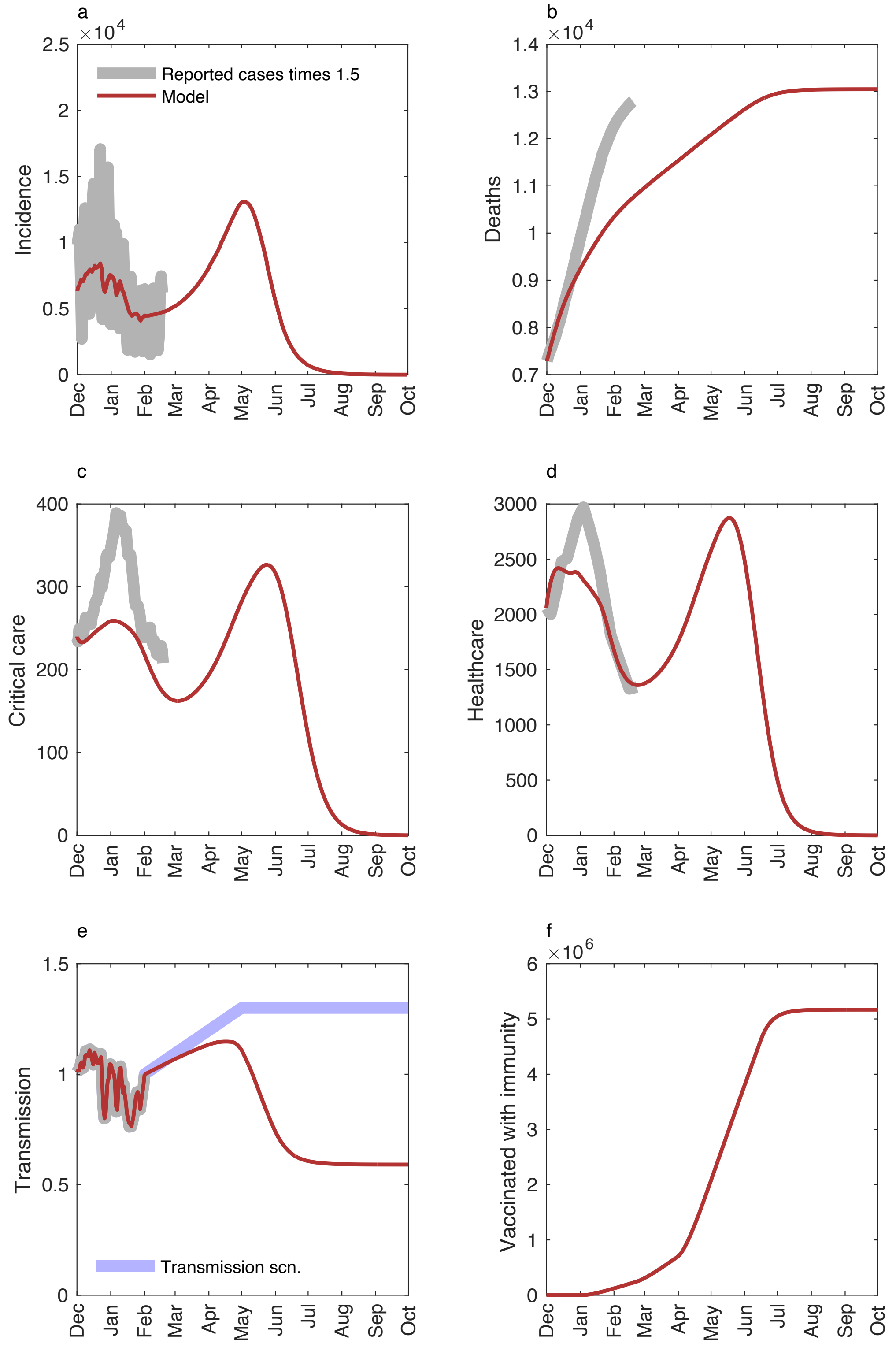


288
289 Figure S 13. Model dynamics for Model scenario 6. Upper left: number of infected, upper right: cumulative
290 deaths, mid left: patients in health care, mid right: patients in critical care, lower left: transmission scenario
291 (blue) and R(t) (red), lower right: number of vaccinated with immunity. See text for additional information.

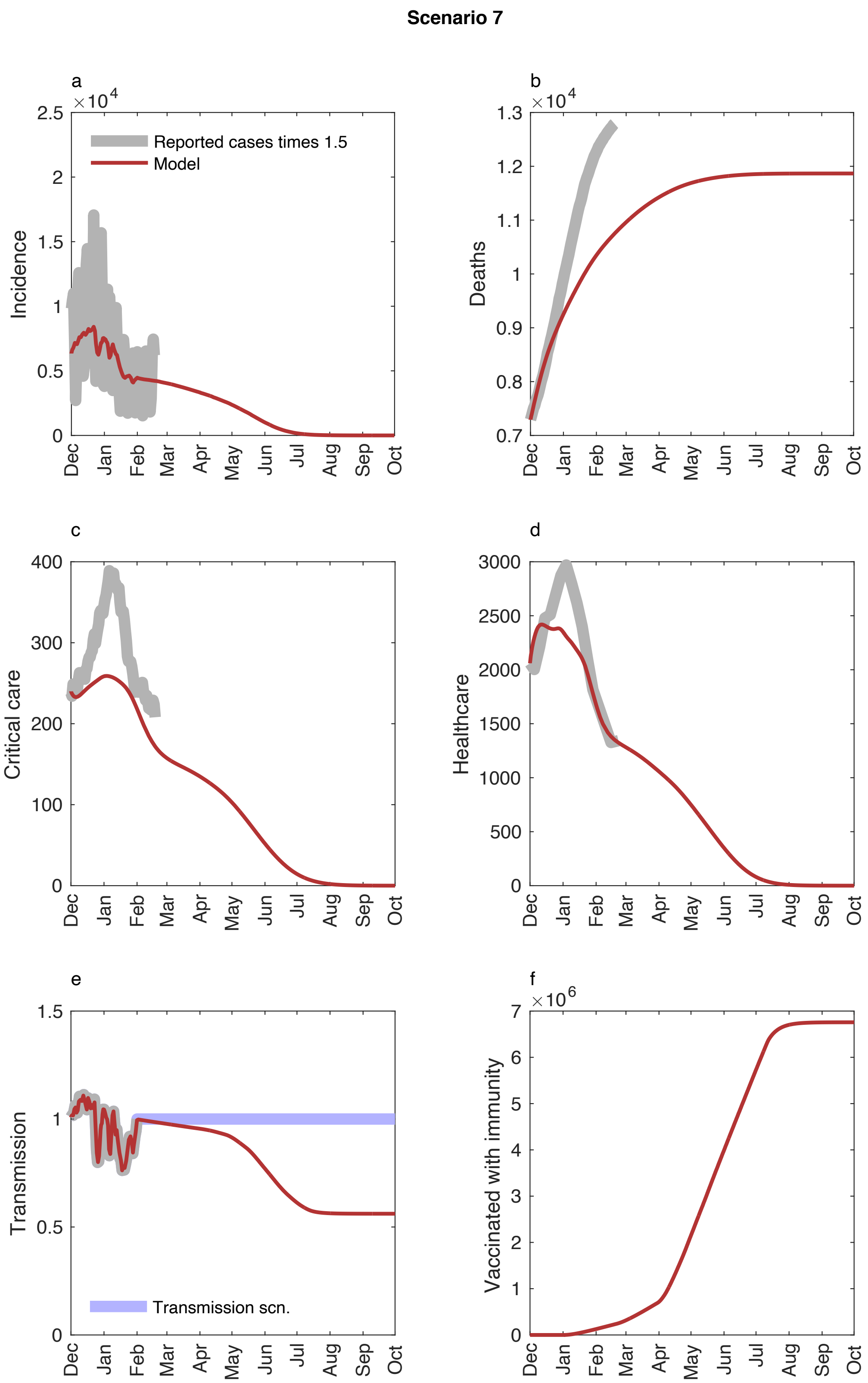


Figure S 14. Model dynamics for Model scenario 7. Upper left: number of infected, upper right: cumulative deaths, mid left: patients in health care, mid right: patients in critical care, lower left: transmission scenario (blue) and R(t) (red), lower right: number of vaccinated with immunity. See text for additional information.

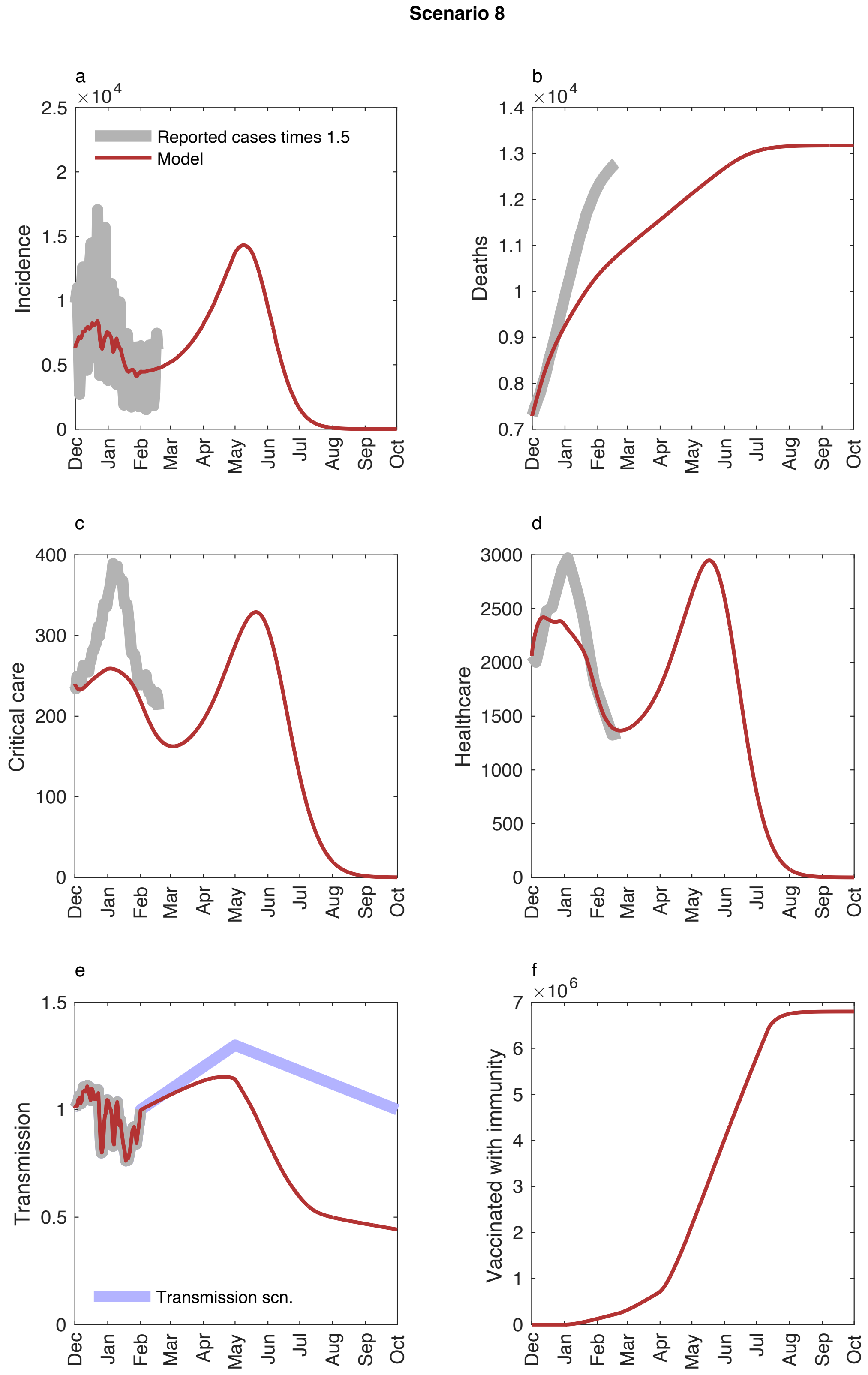


Figure S 15. Model dynamics for Model scenario 8. Upper left: number of infected, upper right: cumulative deaths, mid left: patients in health care, mid right: patients in critical care, lower left: transmission scenario (blue) and R(t) (red), lower right: number of vaccinated with immunity. See text for additional information.

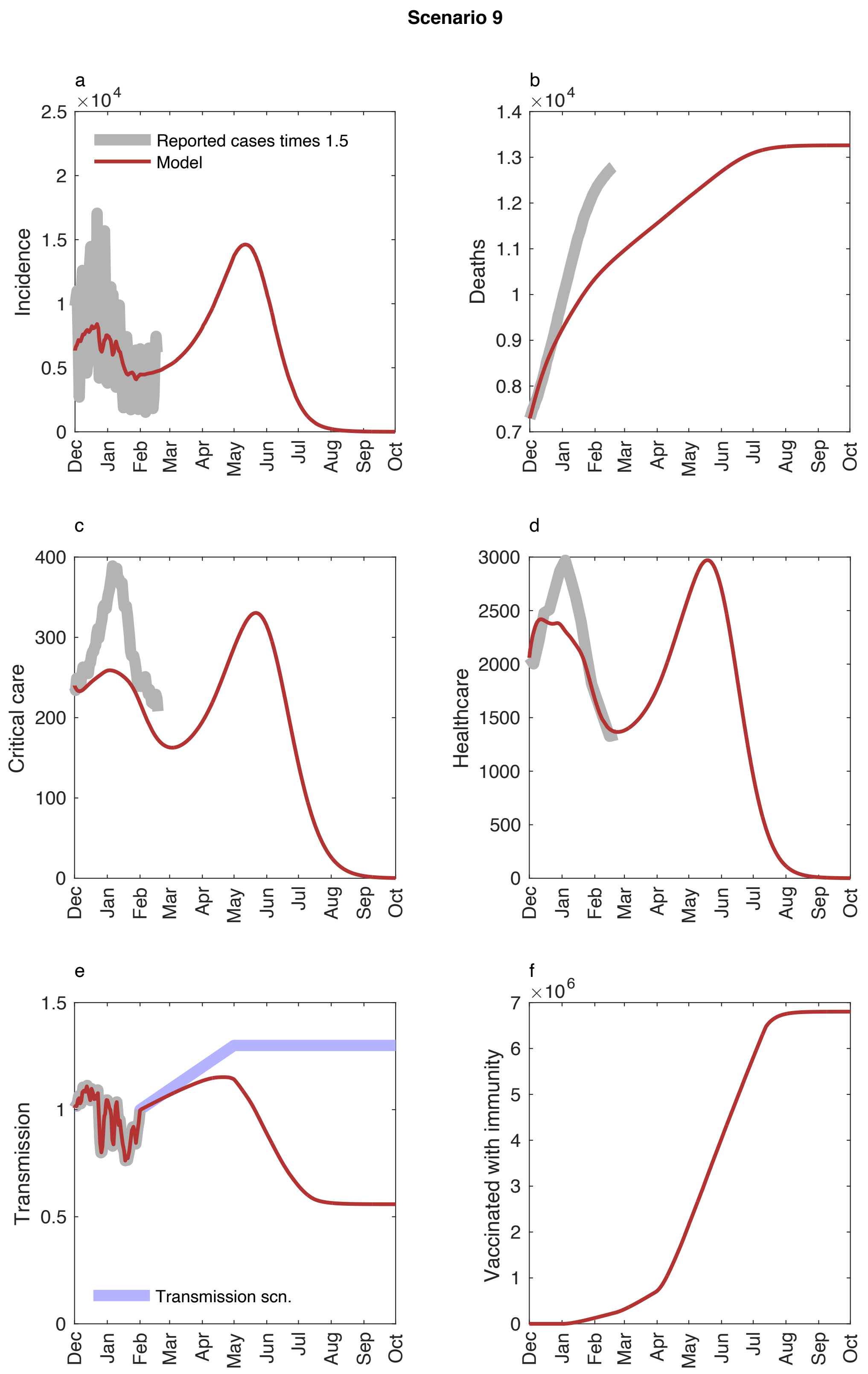


Figure S 16. Model dynamics for Model scenario 9. Upper left: number of infected, upper right: cumulative deaths, mid left: patients in health care, mid right: patients in critical care, lower left: transmission scenario (blue) and R(t) (red), lower right: number of vaccinated with immunity. See text for additional information.

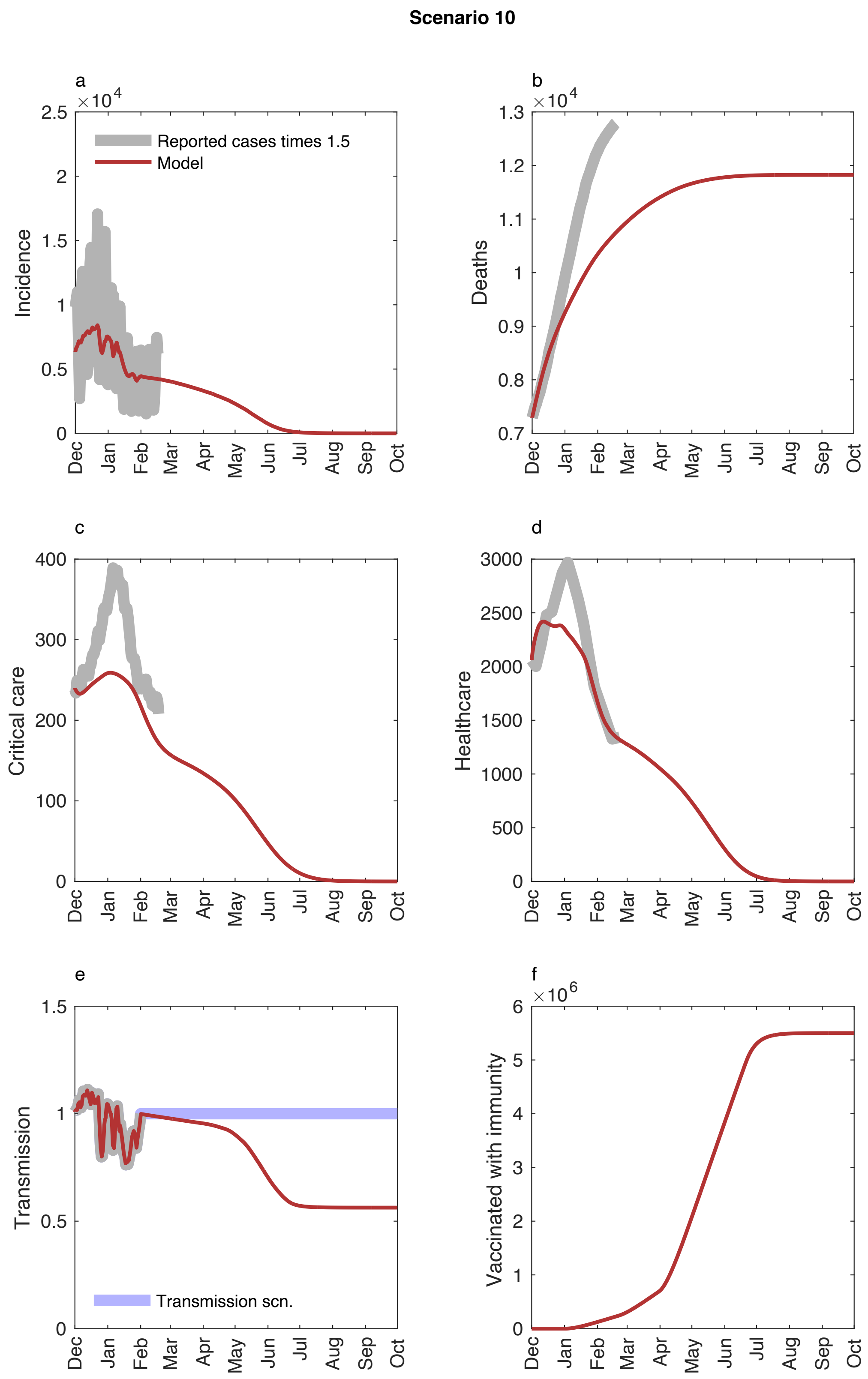


Figure S 17. Model dynamics for Model scenario 10. Upper left: number of infected, upper right: cumulative deaths, mid left: patients in health care, mid right: patients in critical care, lower left: transmission scenario (blue) and R(t) (red), lower right: number of vaccinated with immunity. See text for additional information.

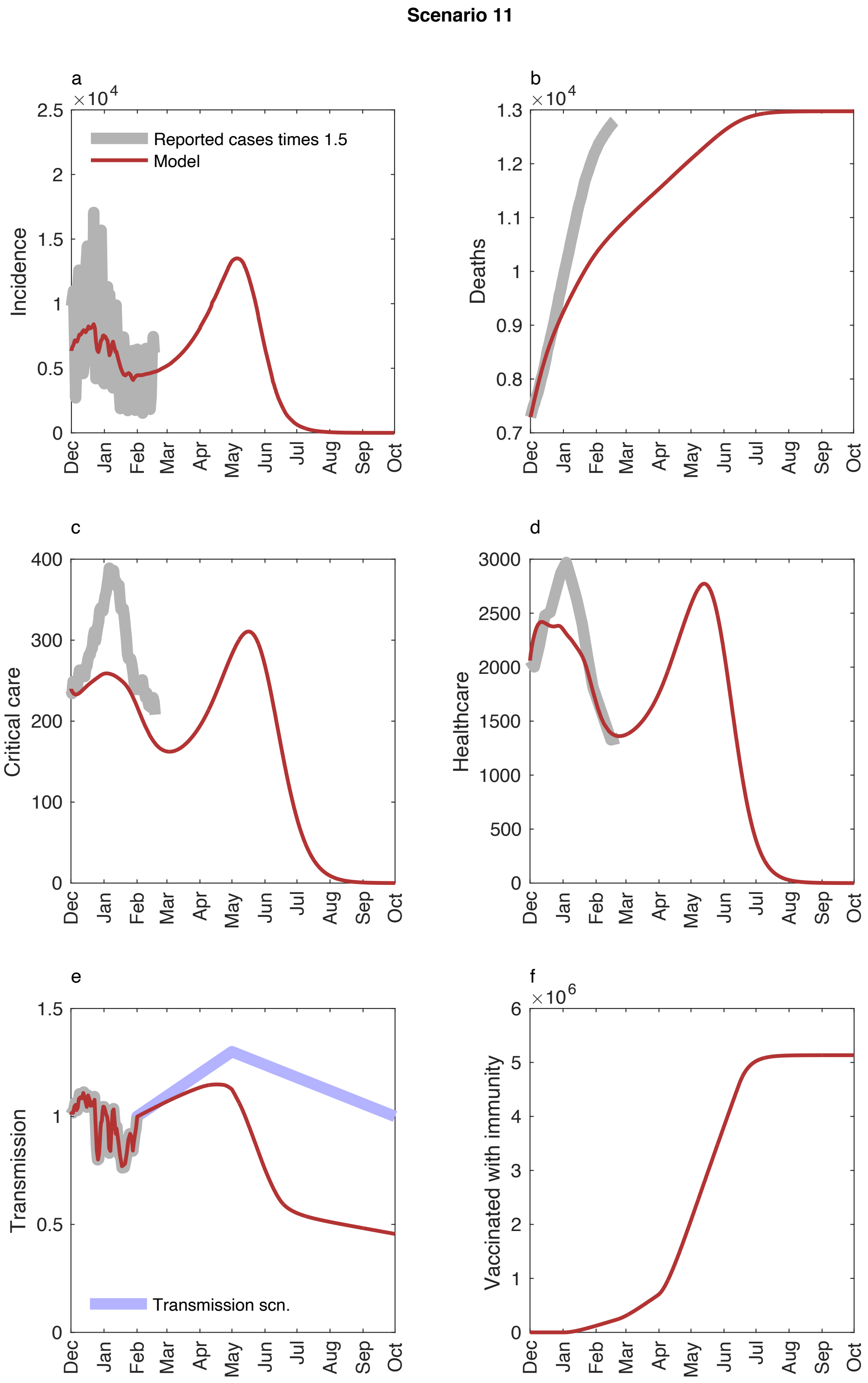


Figure S 18. Model dynamics for Model scenario 11. Upper left: number of infected, upper right: cumulative deaths, mid left: patients in health care, mid right: patients in critical care, lower left: transmission scenario (blue) and R(t) (red), lower right: number of vaccinated with immunity. See text for additional information.

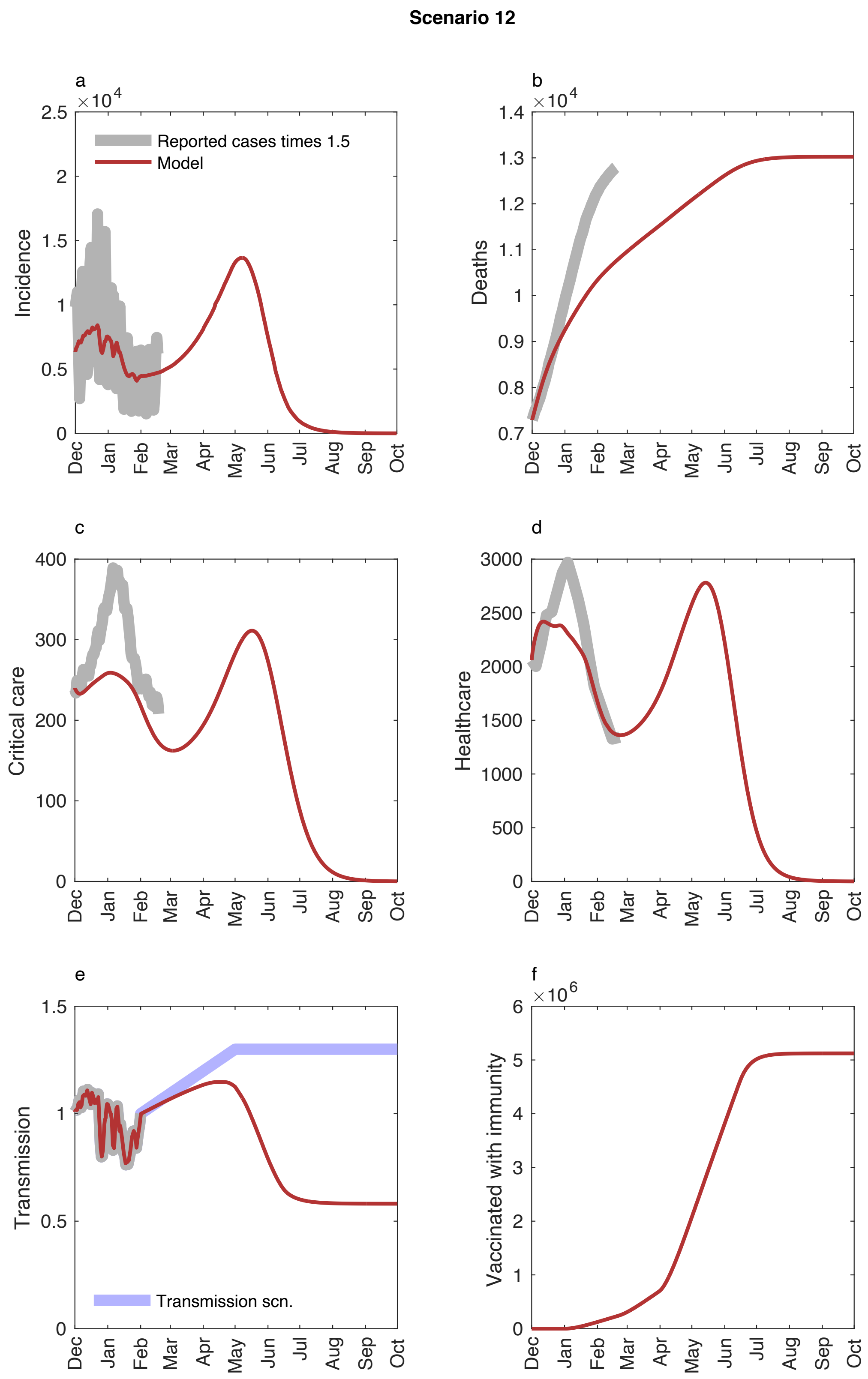


Figure S 19. Model dynamics for Model scenario 12. Upper left: number of infected, upper right: cumulative deaths, mid left: patients in health care, mid right: patients in critical care, lower left: transmission scenario (blue) and R(t) (red), lower right: number of vaccinated with immunity. See text for additional information.

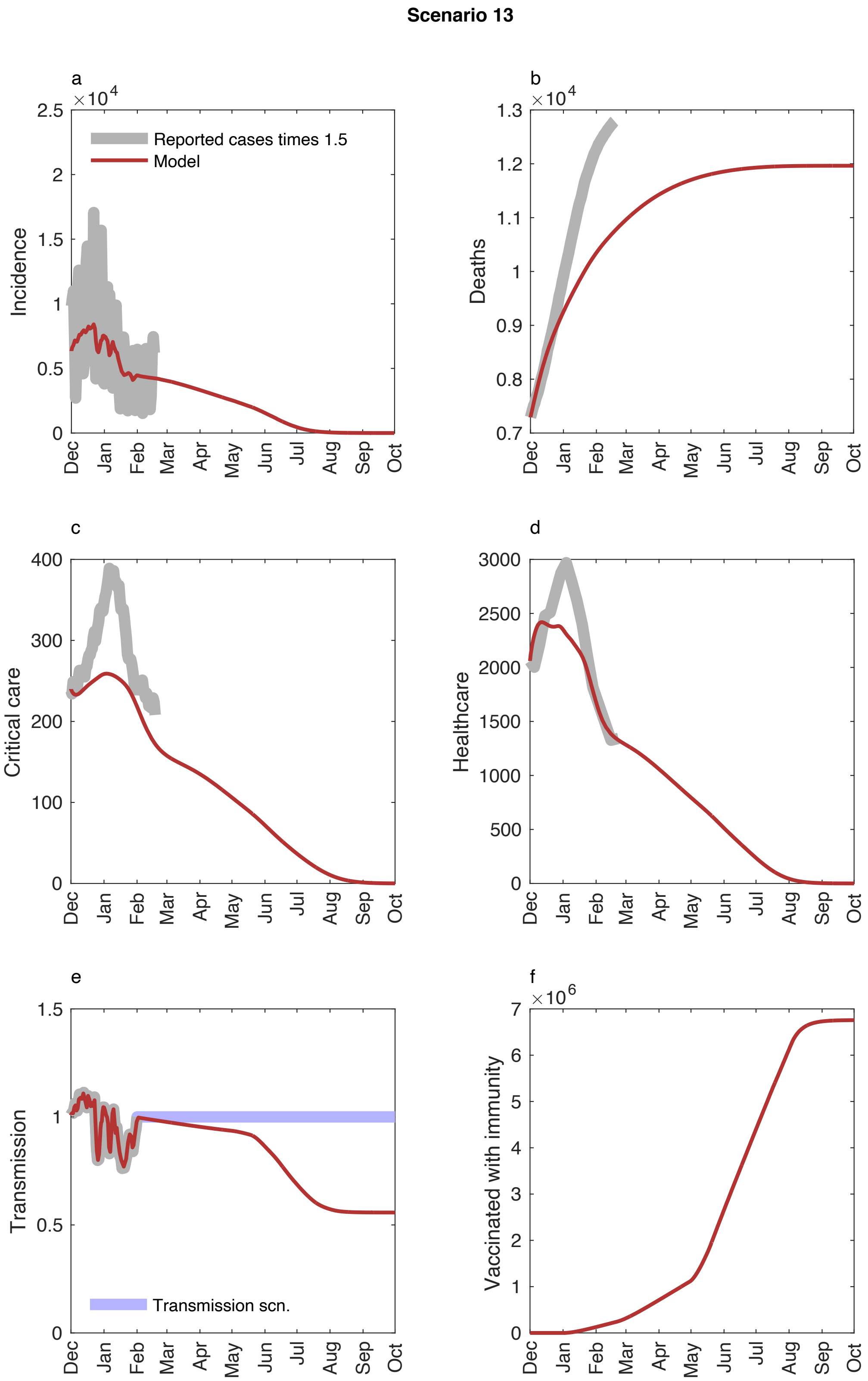


Figure S 20. Model dynamics for Model scenario 13. Upper left: number of infected, upper right: cumulative deaths, mid left: patients in health care, mid right: patients in critical care, lower left: transmission scenario (blue) and R(t) (red), lower right: number of vaccinated with immunity. See text for additional information.

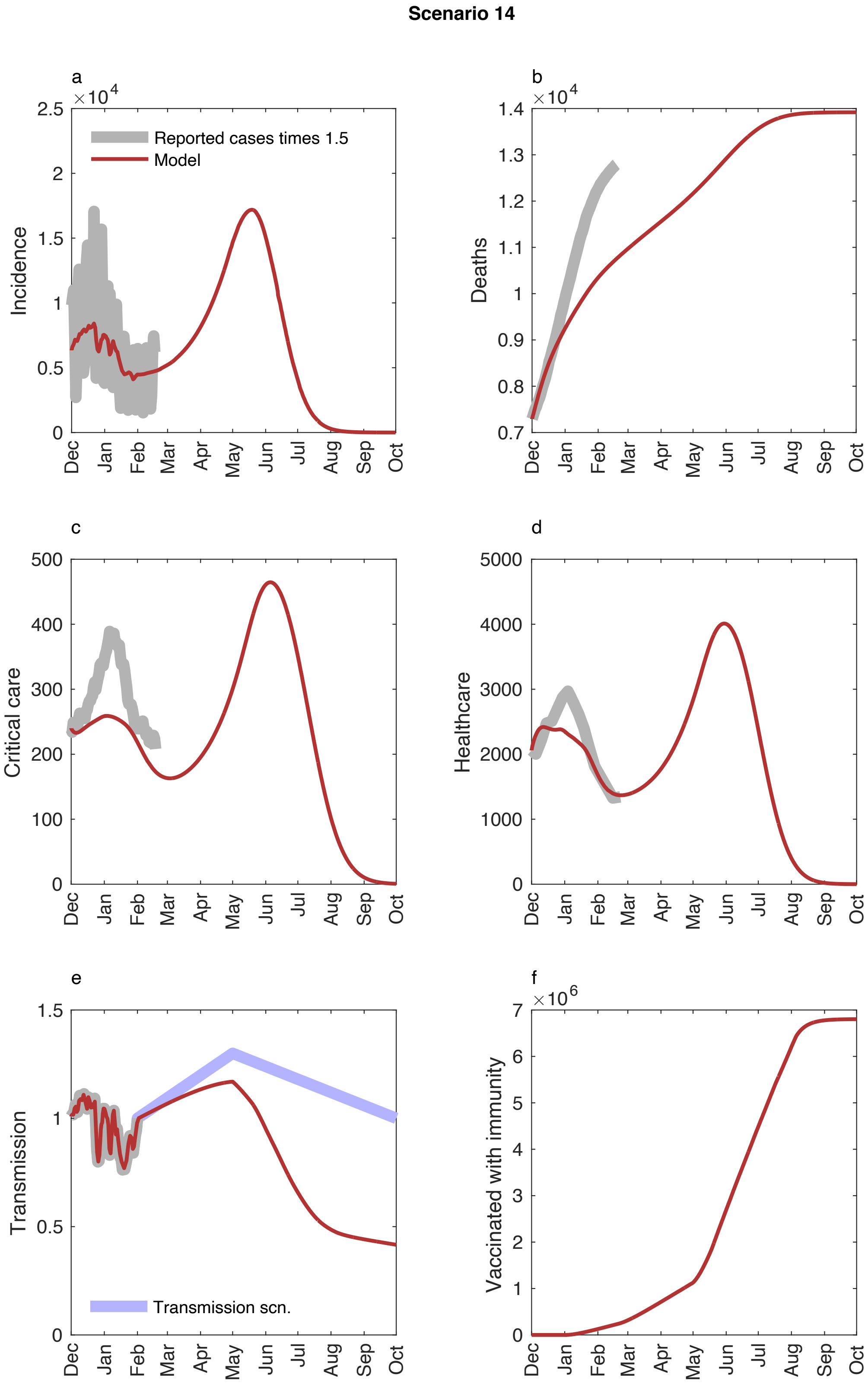


Figure S 21. Model dynamics for Model scenario 14. Upper left: number of infected, upper right: cumulative deaths, mid left: patients in health care, mid right: patients in critical care, lower left: transmission scenario (blue) and R(t) (red), lower right: number of vaccinated with immunity. See text for additional information.

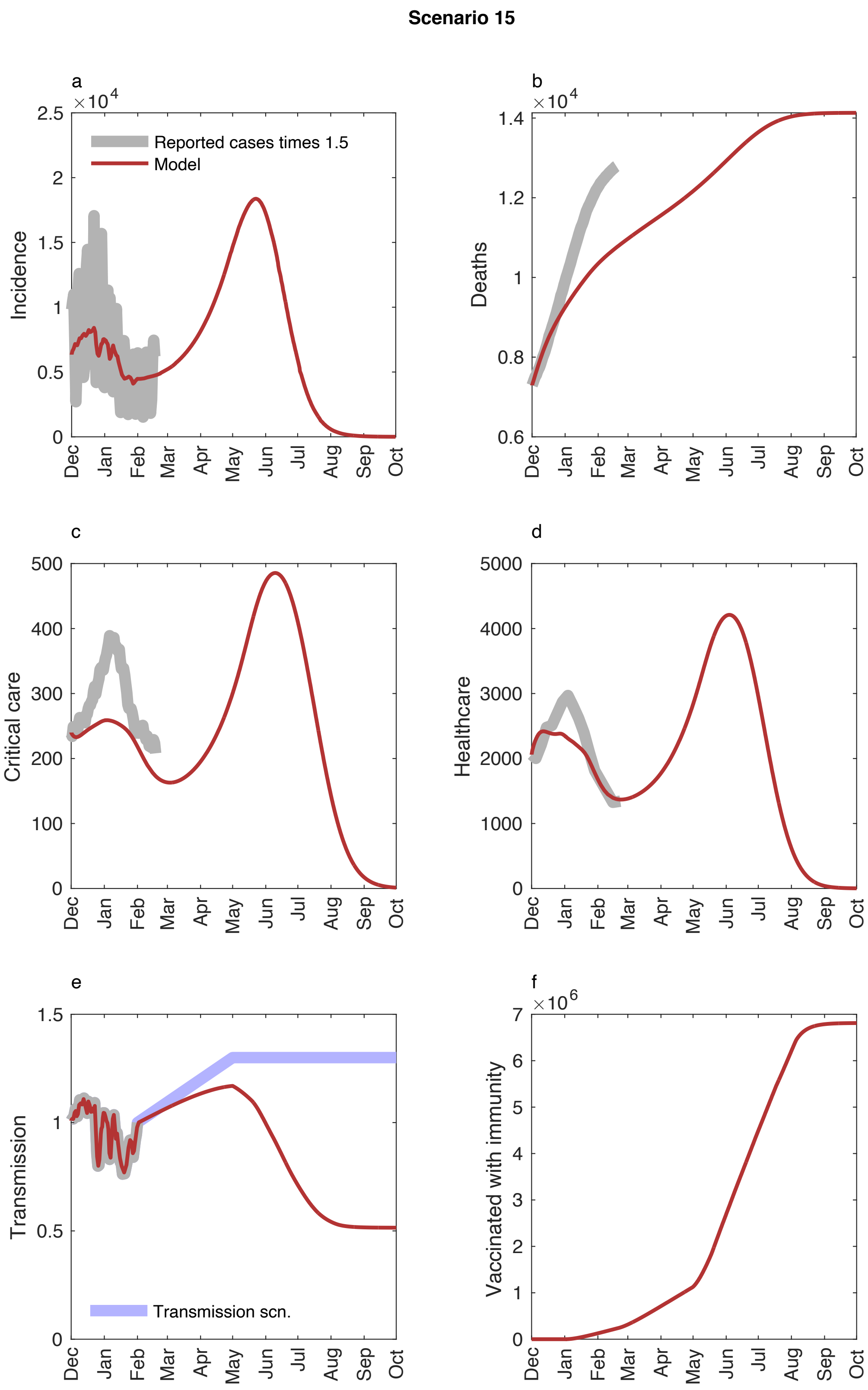


Figure S 22. Model dynamics for Model scenario 15. Upper left: number of infected, upper right: cumulative deaths, mid left: patients in health care, mid right: patients in critical care, lower left: transmission scenario (blue) and R(t) (red), lower right: number of vaccinated with immunity. See text for additional information.

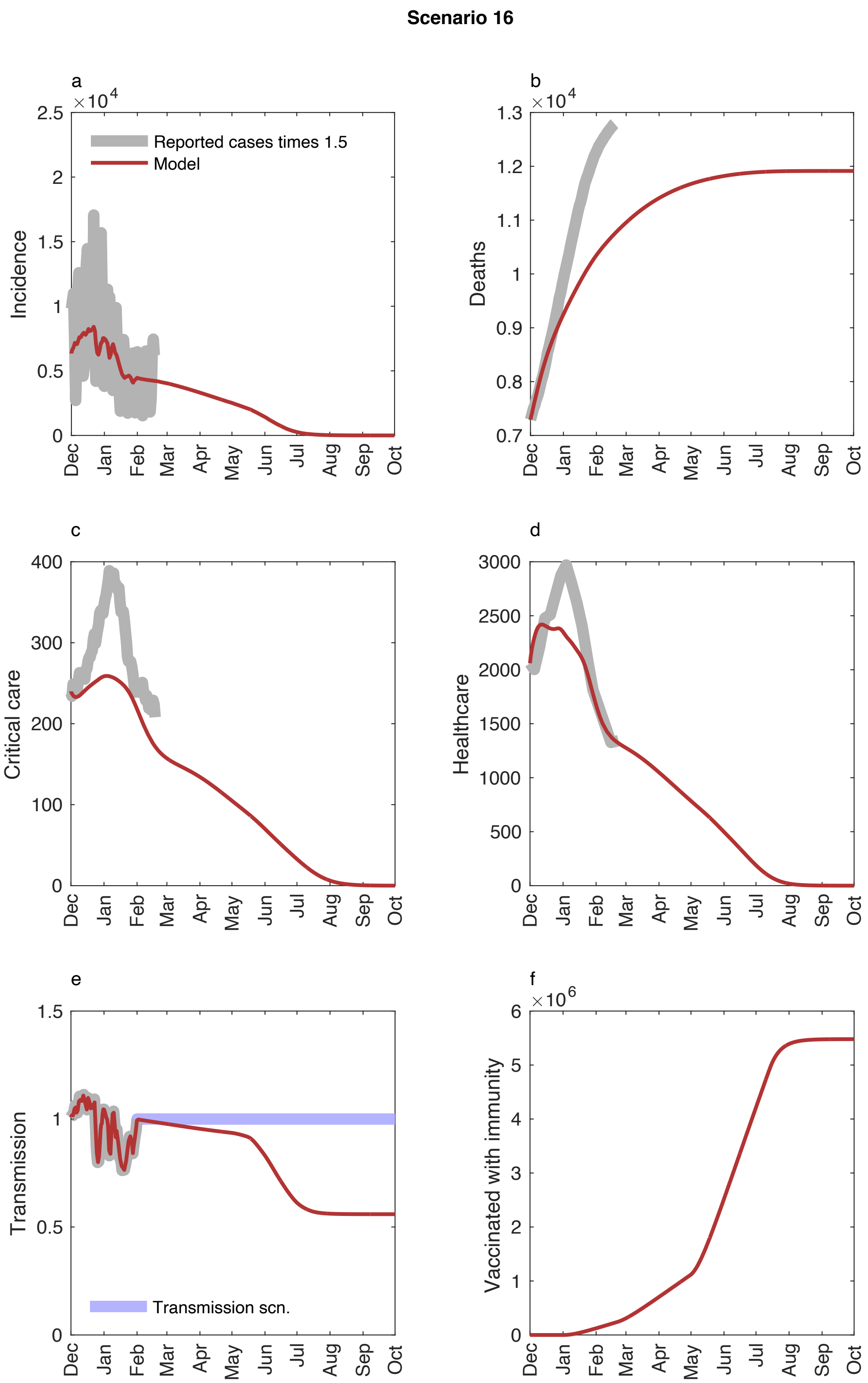


Figure S 23. Model dynamics for Model scenario 16. Upper left: number of infected, upper right: cumulative deaths, mid left: patients in health care, mid right: patients in critical care, lower left: transmission scenario (blue) and R(t) (red), lower right: number of vaccinated with immunity. See text for additional information.

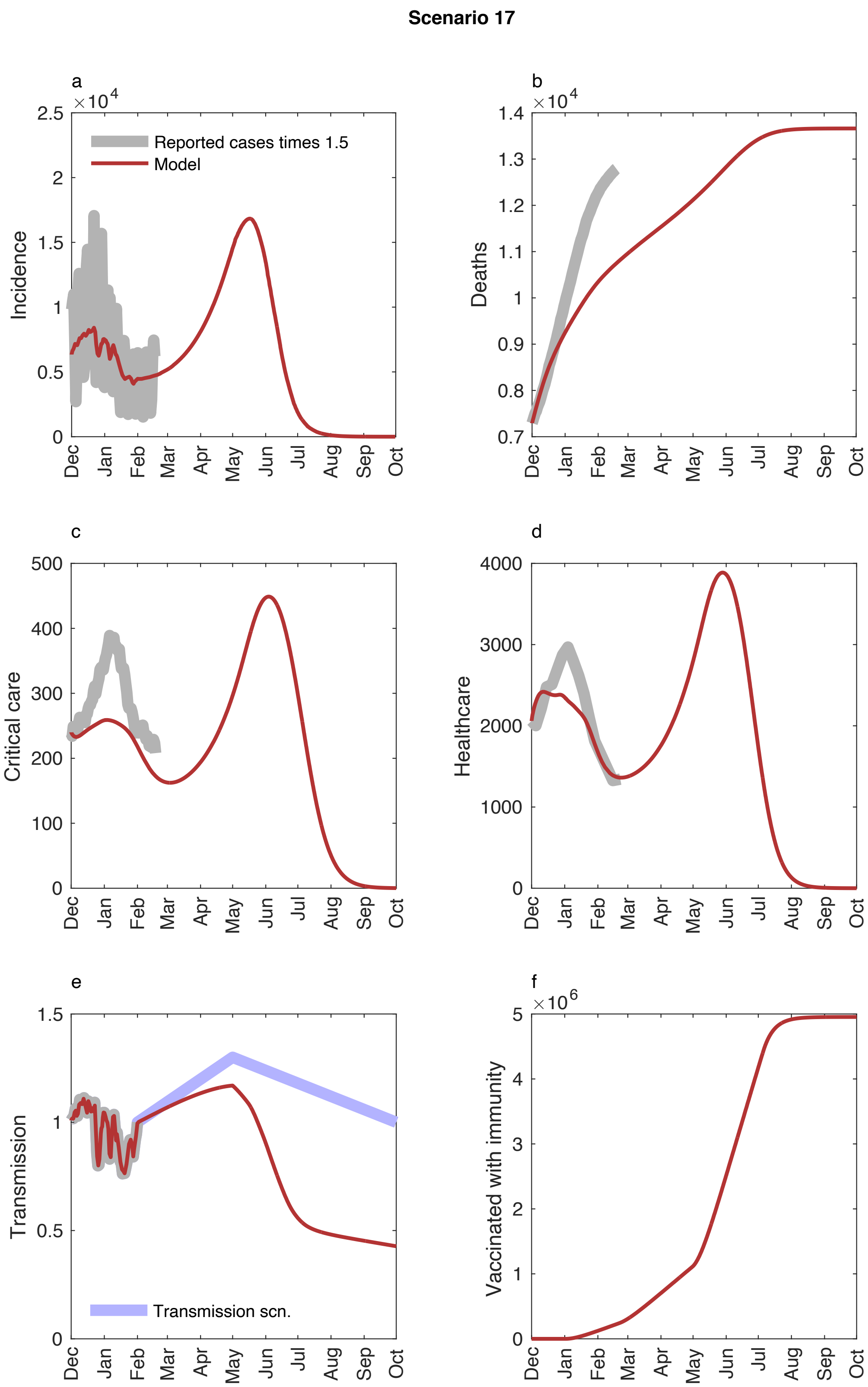


Figure S 24. Model dynamics for Model scenario 17. Upper left: number of infected, upper right: cumulative deaths, mid left: patients in health care, mid right: patients in critical care, lower left: transmission scenario (blue) and R(t) (red), lower right: number of vaccinated with immunity. See text for additional information.

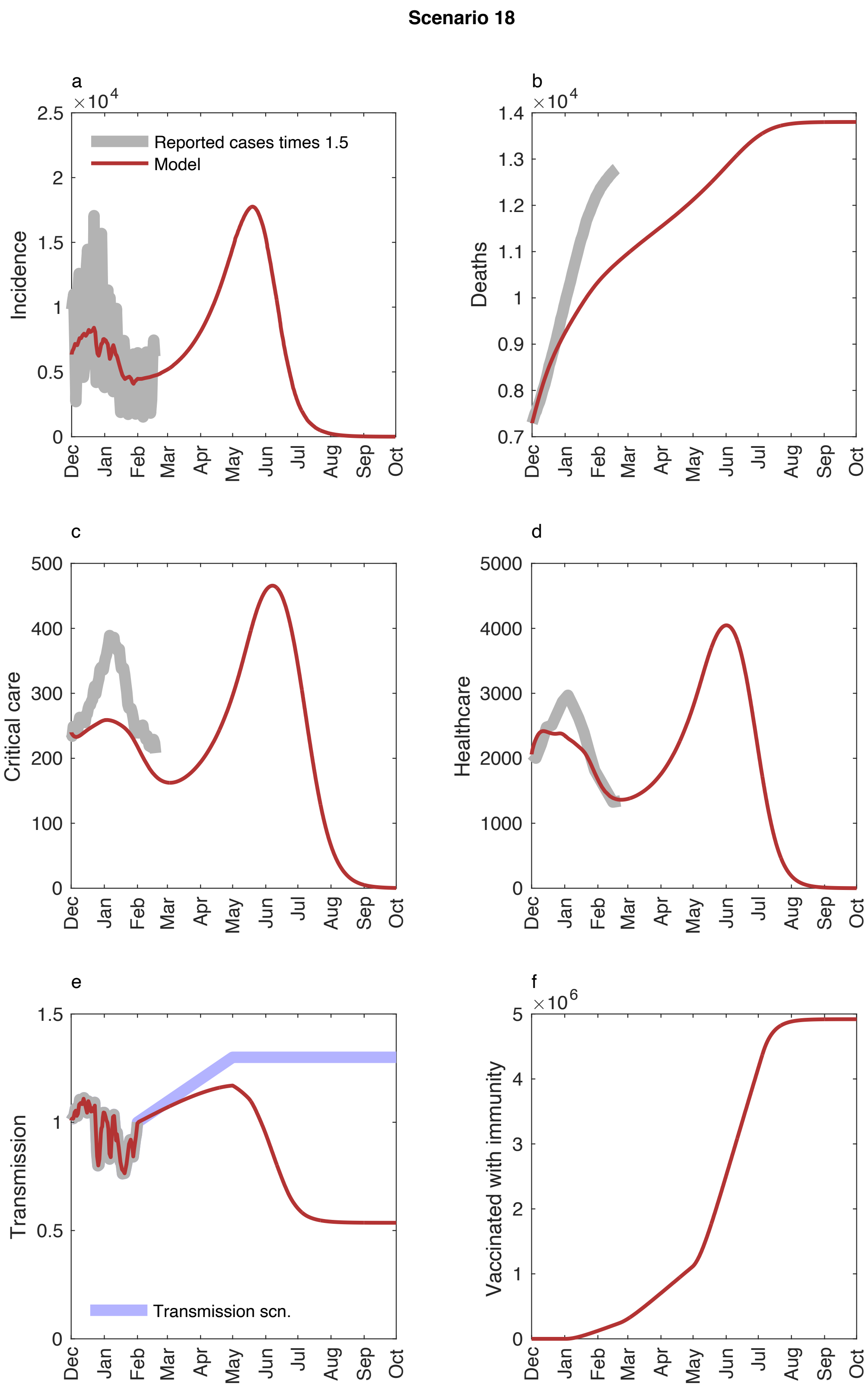


Figure S 25. Model dynamics for Model scenario 18. Upper left: number of infected, upper right: cumulative deaths, mid left: patients in health care, mid right: patients in critical care, lower left: transmission scenario (blue) and R(t) (red), lower right: number of vaccinated with immunity. See text for additional information.

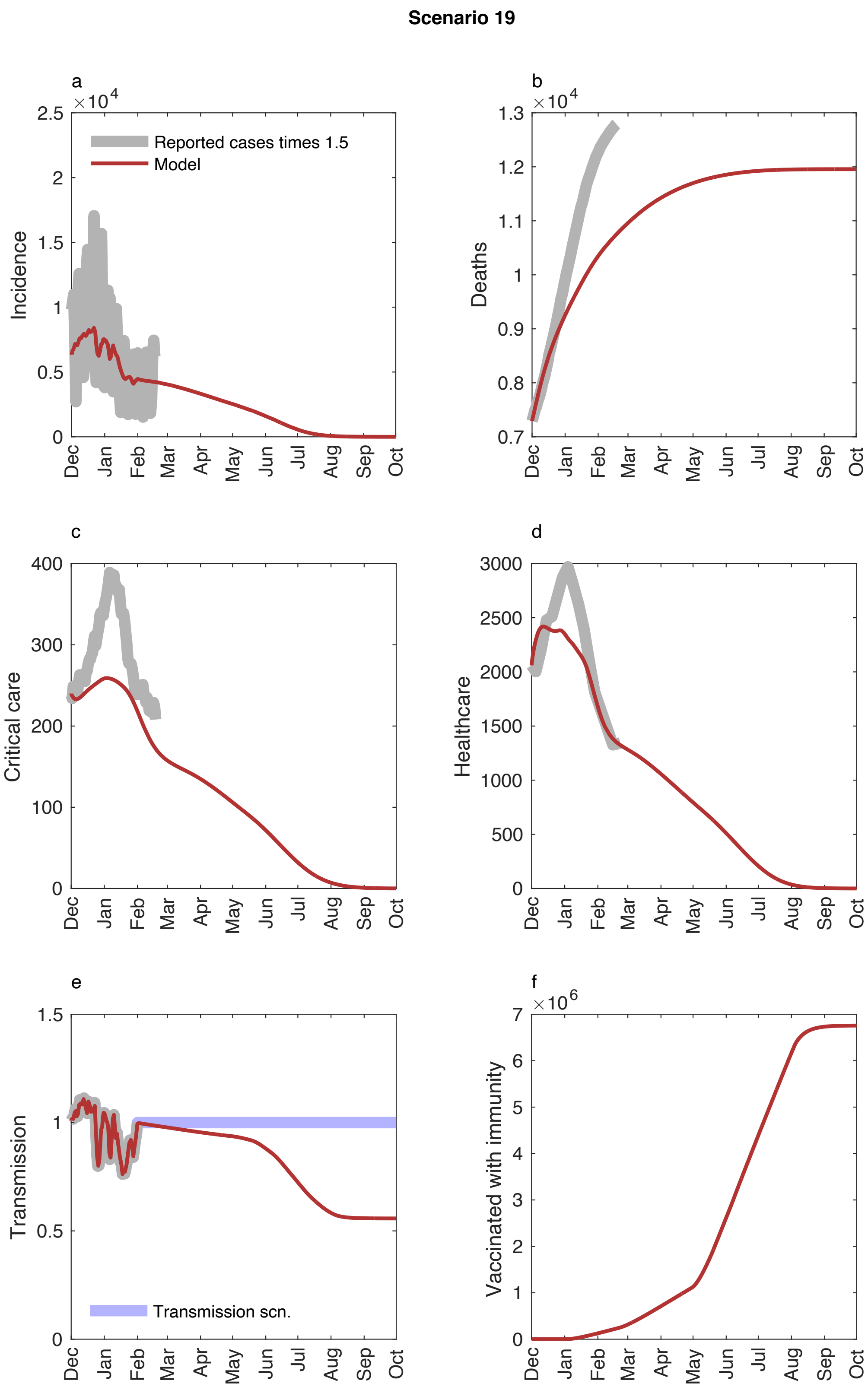


Figure S 26. Model dynamics for Model scenario 19. Upper left: number of infected, upper right: cumulative deaths, mid left: patients in health care, mid right: patients in critical care, lower left: transmission scenario (blue) and R(t) (red), lower right: number of vaccinated with immunity. See text for additional information.

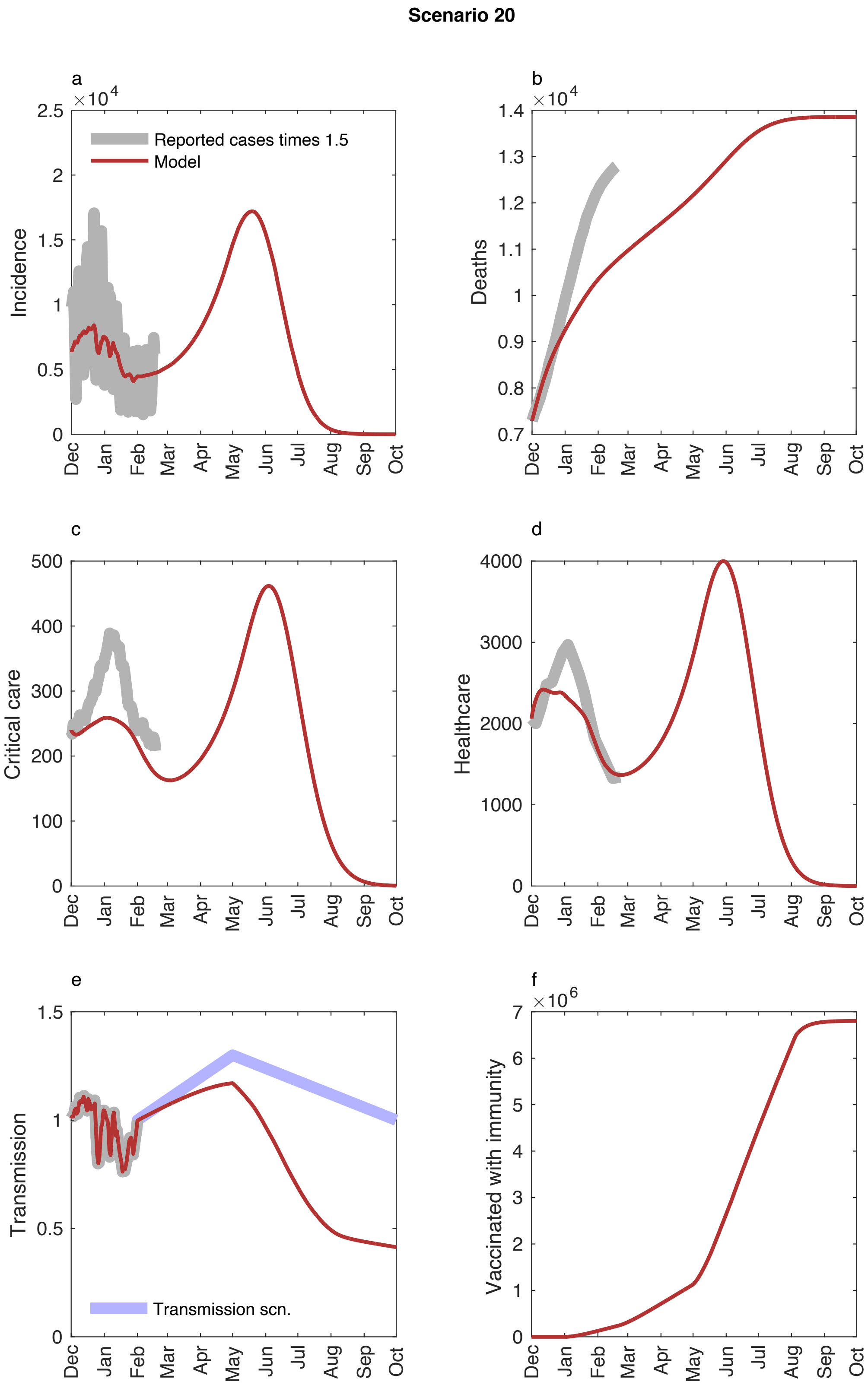


Figure S 27. Model dynamics for Model scenario 20. Upper left: number of infected, upper right: cumulative deaths, mid left: patients in health care, mid right: patients in critical care, lower left: transmission scenario (blue) and R(t) (red), lower right: number of vaccinated with immunity. See text for additional information.

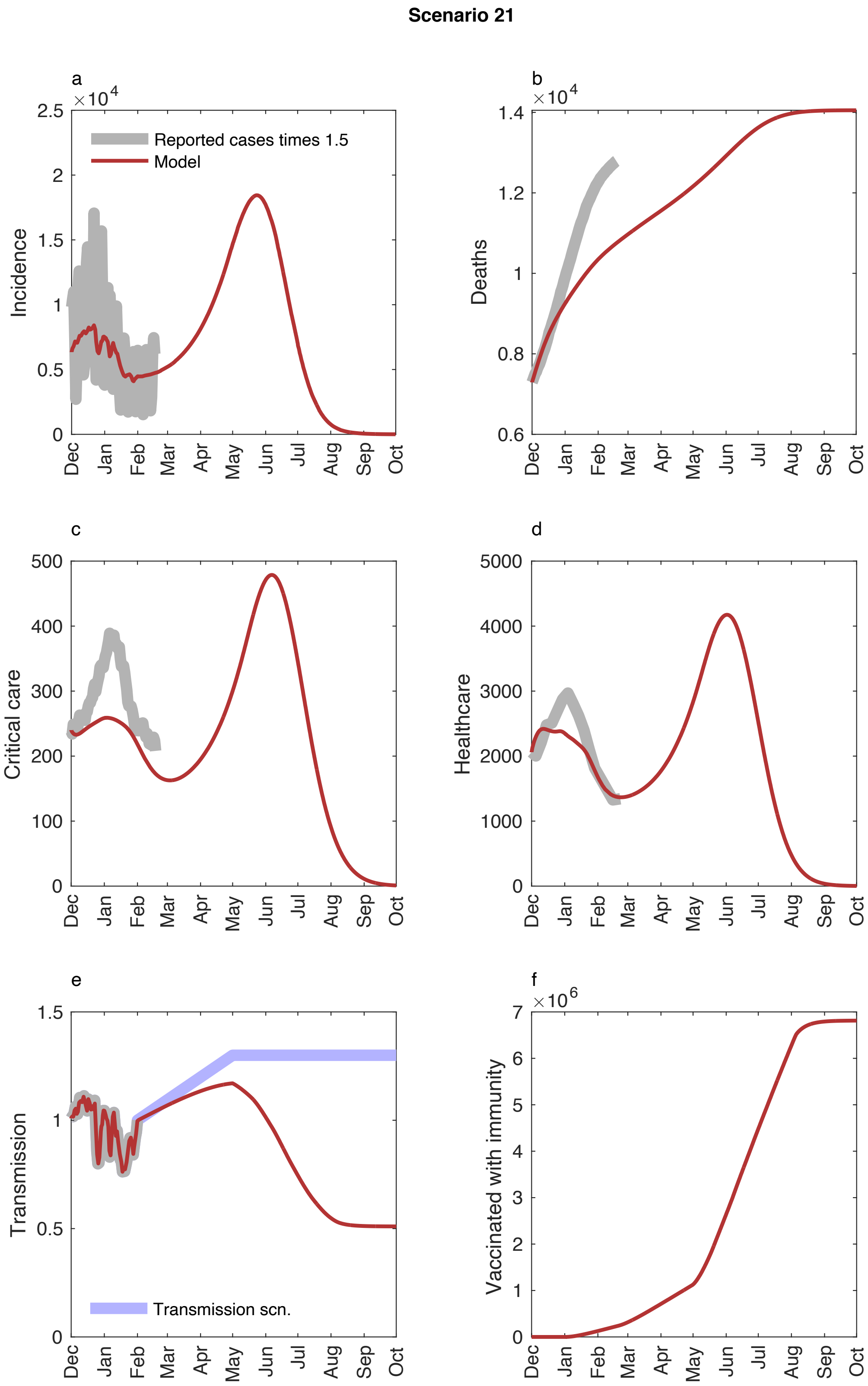


Figure S 28. Model dynamics for Model scenario 21. Upper left: number of infected, upper right: cumulative deaths, mid left: patients in health care, mid right: patients in critical care, lower left: transmission scenario (blue) and R(t) (red), lower right: number of vaccinated with immunity. See text for additional information.

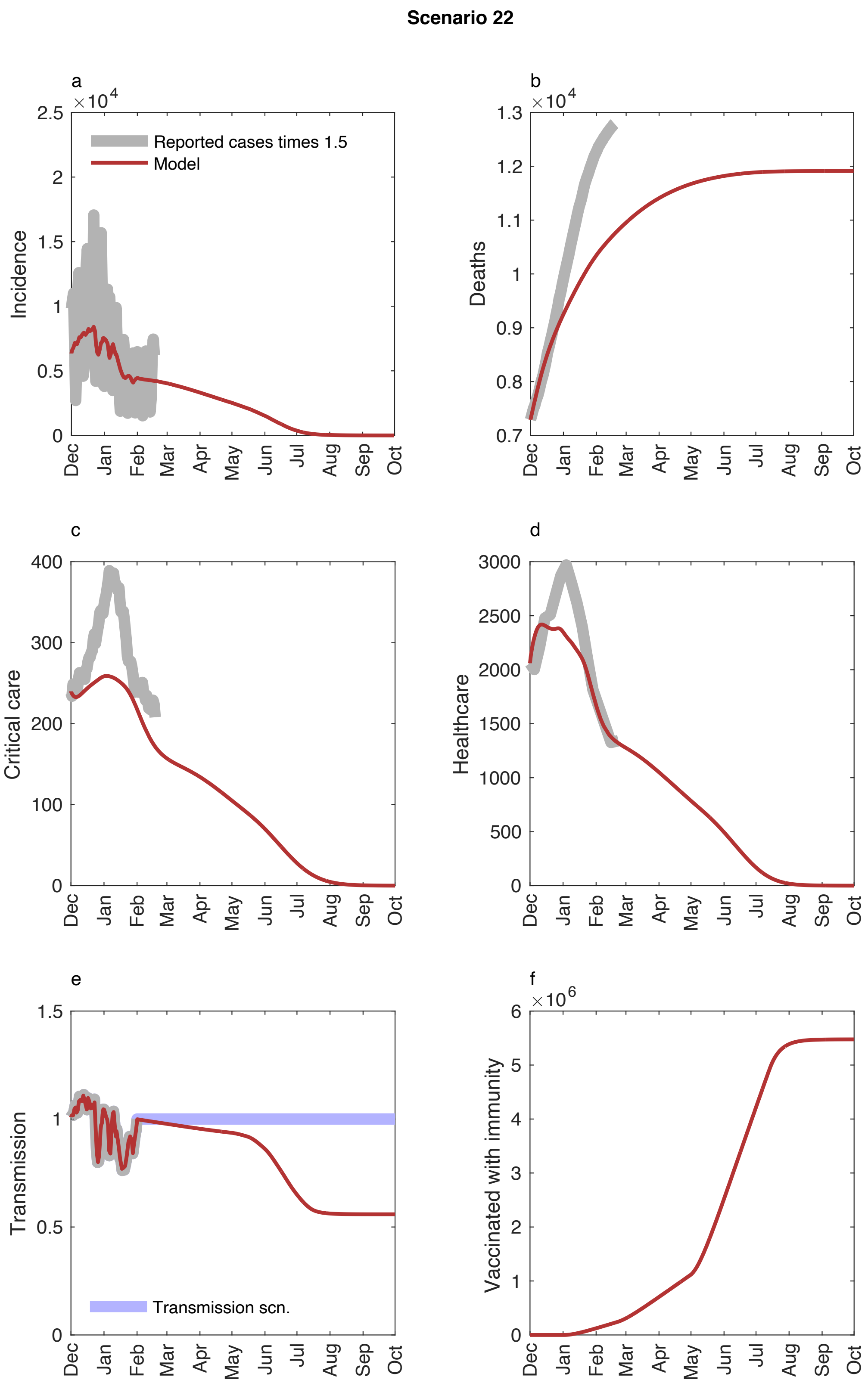


Figure S 29. Model dynamics for Model scenario 22. Upper left: number of infected, upper right: cumulative deaths, mid left: patients in health care, mid right: patients in critical care, lower left: transmission scenario (blue) and R(t) (red), lower right: number of vaccinated with immunity. See text for additional information.

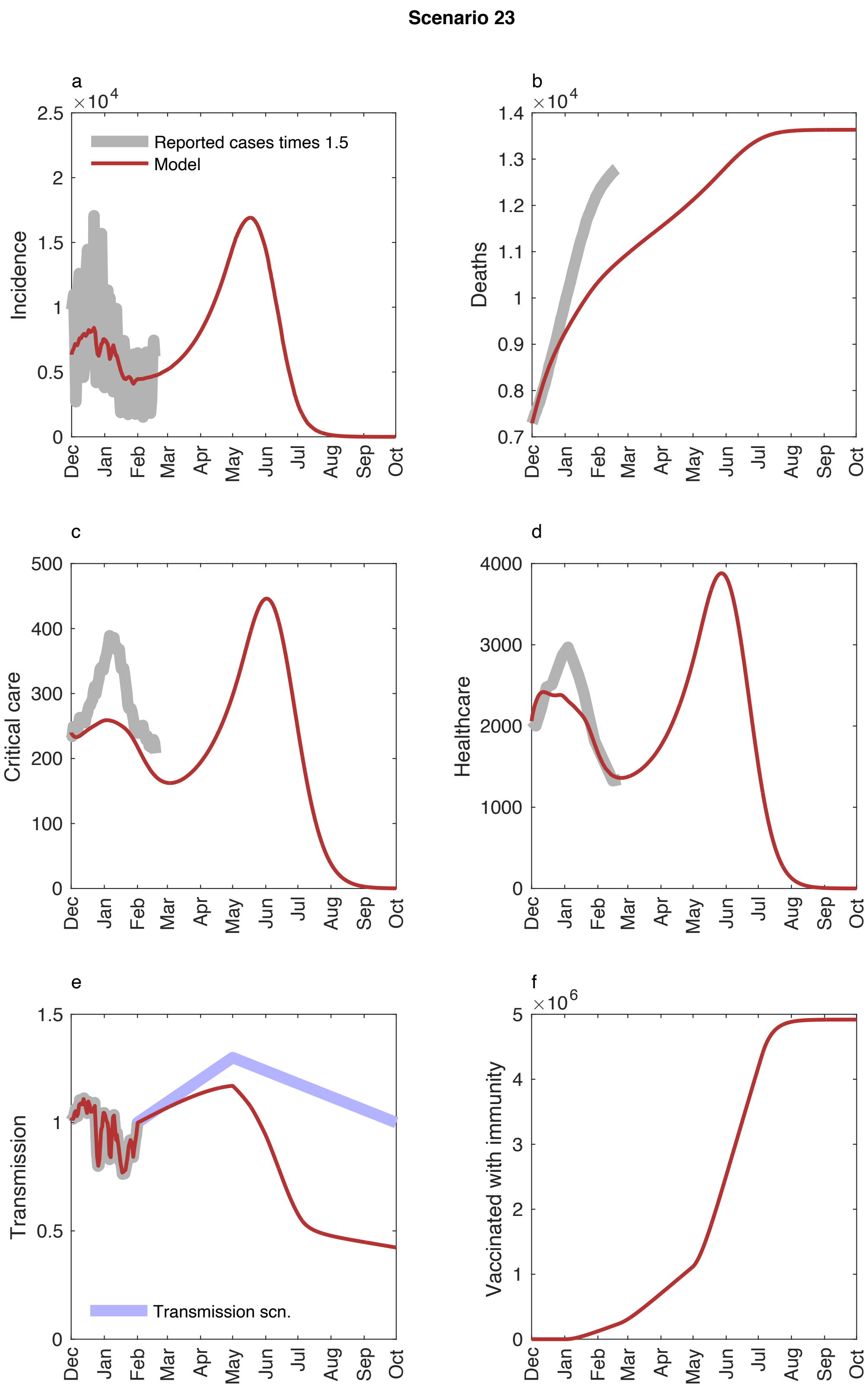


Figure S 30. Model dynamics for Model scenario 23. Upper left: number of infected, upper right: cumulative deaths, mid left: patients in health care, mid right: patients in critical care, lower left: transmission scenario (blue) and R(t) (red), lower right: number of vaccinated with immunity. See text for additional information.

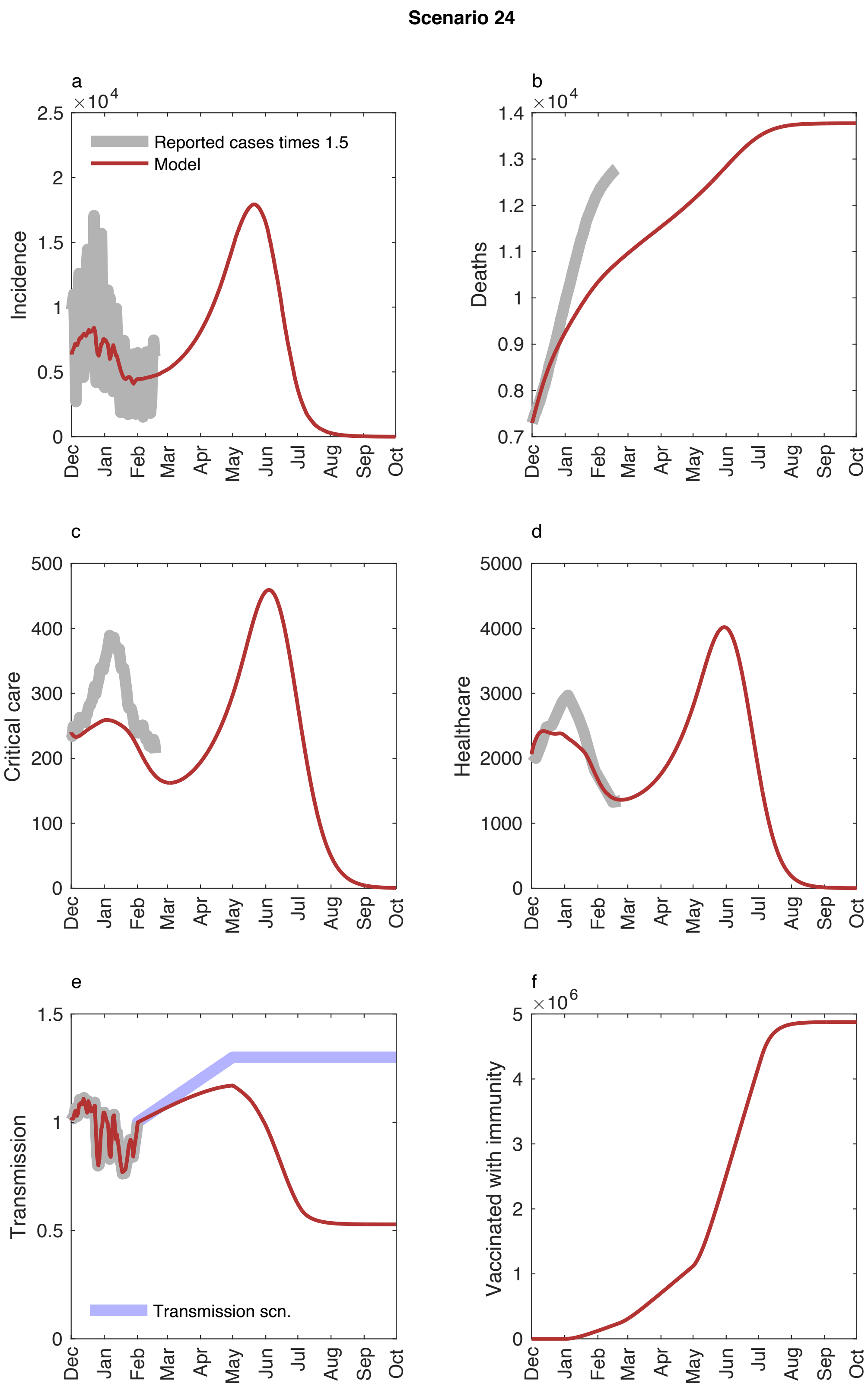


Figure S 31. Model dynamics for Model scenario 24. Upper left: number of infected, upper right: cumulative deaths, mid left: patients in health care, mid right: patients in critical care, lower left: transmission scenario (blue) and R(t) (red), lower right: number of vaccinated with immunity. See text for additional information.

### References

1. Prem K, Cook AR, Jit M. Projecting social contact matrices in 152 countries using contact surveys and demographic data. *PLoS Comput Biol*, 2017, 13(9): e1005697. https://doi.org/10.1371/journal.pcbi.1005697

2. The Public Health Agency of Sweden, https://www.folkhalsomyndigheten.se

3. The  Swedish National Board of Health and Welfare, https://www.socialstyrelsen.se

4. Webpage, www.c19.se

5. Sun K, et al. Transmission heterogeneities, kinetics, and controllability of SARS-CoV-2, *Science*, 2021, 371.6526.

6. Swedish National Board of Health and Welfare, https://www.socialstyrelsen.se/om-socialstyrelsen/pressrum/press/langa-vardtider-pa-sjukhus-for-patienter-med-covid-19/

7. https://lakartidningen.se/aktuellt/nyheter/2020/12/halverad-vardtid-for-iva-patienter-med-covid/

8. https://www.socialstyrelsen.se/om-socialstyrelsen/pressrum/press/halvering-av-andelen-doda-bland-dem-som-sjukhusvardats-for-covid-19/
